# Incisor Extraction in Orthodontics: A Systematic Review and Meta-Analysis of Clinical Outcomes and Biomechanics

**DOI:** 10.64898/2026.03.23.26349102

**Authors:** Maen Mahfouz, Eman Alzaben

## Abstract

**Background:** Incisor extraction represents a strategic yet underutilized orthodontic treatment modality for managing anterior discrepancies. Despite its clinical relevance, the evidence base has not been systematically synthesized with meta-analytic techniques.

**Objective:** To systematically review and meta-analyze the evidence on incisor extraction in orthodontic treatment, evaluating clinical outcomes and biomechanical efficacy in both maxillary and mandibular arches.

**Methods:** A comprehensive search of open-access electronic databases (PubMed, LILACS, SciELO, Google Scholar, DOAJ, OpenGrey) and orthodontic journal archives was conducted from inception to January 11, 2026 following PRISMA guidelines. Eligible studies included randomized controlled trials, prospective cohort studies, and retrospective cohort studies with ≥10 patients reporting quantitative outcomes following incisor extraction or incisor movement with premolar extraction. Primary outcomes included space closure efficiency, incisor position changes, root resorption, and stability. Risk of bias was assessed using ROBINS-I for observational studies and Cochrane RoB 2.0 for RCTs. Certainty of evidence was evaluated using GRADE.

**Results:** From 1,842 identified records, 20 primary studies met inclusion criteria (4 RCTs, 16 observational studies), comprising 1,347 patients. Sixteen studies provided data for meta-analysis. With moderate-certainty evidence, mandibular incisor extraction (8 studies, n=412) demonstrated mean space closure of 5.2 mm (95% CI 4.8 to 5.6 mm, I²=34%) with favorable long-term stability (mean irregularity increase 0.3 mm, 95% CI 0.1 to 0.5 mm, I²=28%). Low-certainty evidence indicates clear aligner accuracy is limited to 78.9% of predicted incisor tip movement (3 studies, n=187, 95% CI 72.3 to 85.5%, I²=41%); these findings may not reflect newer generation aligner systems. Low-certainty evidence suggests maxillary incisor movement following premolar extraction (6 studies using tomographic imaging, n=387) results in palatal bone resorption (mean −0.43 mm, 95% CI −0.62 to −0.24 mm, I²=52%), with greater effects in adults versus adolescents (mean difference 0.31 mm, p = 0.02); although statistically significant, the magnitude may be clinically negligible in patients with adequate baseline alveolar thickness. Moderate-certainty evidence indicates en-masse retraction results in faster space closure than two-step retraction (4 RCTs, n=214, mean −4.2 months, 95% CI −5.8 to −2.6 months). Moderate-certainty evidence shows root resorption incidence is 12.4% (95% CI 8.7 to 16.1%), with subgroup analysis: >2 mm threshold 13.2% (7 studies), ≥¼ root length threshold 11.4% (5 studies). Low-certainty evidence suggests extraction versus non-extraction comparisons (4 studies, n=326) show no significant differences in relapse.

**Conclusions:** Mandibular incisor extraction demonstrates favorable long-term stability with minimal profile changes but requires recognition of clear aligner accuracy limitations. Maxillary incisor movement carries risks including palatal bone resorption, particularly in adults, though the clinical significance may vary with baseline alveolar thickness. En-masse retraction results in faster space closure with comparable root resorption risk. Treatment decisions should consider patient-specific factors including age, alveolar bone morphology, malocclusion pattern, and appliance selection.

## INTRODUCTION

Incisor extraction represents a strategic yet underutilized orthodontic treatment modality that offers distinct advantages in carefully selected clinical scenarios. Unlike premolar extractions, which affect the entire dental arch and often necessitate significant retraction of the anterior segment, incisor extraction offers a localized space-management solution that can address anterior discrepancies with minimal impact on the facial profile [1].

The decision to extract an incisor is driven by multiple factors: the need to resolve severe localized crowding, correction of tooth size discrepancies (Bolton analysis) [2], camouflage of specific skeletal patterns, or management of compromised teeth due to trauma, ectopic eruption, or developmental anomalies [3]. Despite its clinical relevance, the evidence base for incisor extraction has not been systematically synthesized with meta-analytic techniques.

Incisor extraction accounts for approximately 2–6% of extraction protocols in survey-based orthodontic cohorts [4,5]. Mandibular incisor extraction is substantially more common than maxillary incisor extraction, reflecting its lower esthetic risk, more predictable biomechanical outcomes, and well-documented indications for managing lower anterior crowding and mild Class III malocclusion [6]. The relative rarity of this procedure underscores the importance of evidence-based guidelines to inform clinical decision-making.

Several controversies warrant systematic investigation. First, stability concerns regarding long-term alignment following single-incisor extraction have been debated [7,8]. Second, the risk of open interproximal embrasures (“black triangles”) presents significant esthetic implications [9]. Third, the impact of technological advances—particularly clear aligner therapy—on treatment accuracy requires quantification [10]. Fourth, bone remodeling patterns following incisor movement, especially in the maxillary arch, remain incompletely characterized [11]. Fifth, the comparative efficacy of en-masse versus two-step retraction techniques continues to be debated [12].

Previous narrative reviews have provided valuable clinical guidance but lack quantitative synthesis. The emergence of high-quality randomized controlled trials and tomographic imaging studies in recent years enables meta-analytic investigation. Therefore, the objective of this systematic review and meta-analysis was to evaluate the clinical efficacy and stability of mandibular incisor extraction, quantify bone remodeling patterns following maxillary incisor movement, compare en-masse versus two-step retraction biomechanics, assess the accuracy of clear aligner therapy in incisor extraction cases, investigate root resorption incidence and risk factors, and compare extraction versus non-extraction outcomes for stability.

## MATERIAL AND METHODS

This systematic review and meta-analysis was conducted following the Preferred Reporting Items for Systematic Reviews and Meta-Analyses (PRISMA) 2020 guidelines [13]. A formal protocol was not prospectively registered; however, the methodology was predefined and strictly adhered to PRISMA 2020 guidelines to minimize bias. A detailed protocol is available from the corresponding author upon reasonable request.

### Eligibility Criteria (PICOS Framework)

| Component | Inclusion Criteria | Exclusion Criteria |
| --- | --- | --- |
| **Population** | Human patients undergoing orthodontic treatment involving incisor extraction or incisor movement following premolar extraction | Animal studies, syndromic patients, cleft lip/palate, craniofacial anomalies |
| **Intervention** | Orthodontic treatment with incisor extraction (mandibular or maxillary) or incisor retraction following premolar extraction | Non-extraction treatments, premolar extraction without incisor analysis |
| **Comparison** | Alternative extraction protocols, non-extraction controls, different retraction techniques, different appliance types | No comparison group (for comparative analyses) |
| **Outcomes** | Space closure amount/efficiency, incisor position changes, root resorption, bone thickness changes, stability/relapse, treatment duration | Qualitative descriptions only, no quantitative outcome data |
| **Study Design** | Randomized controlled trials, prospective cohort studies, retrospective cohort studies with $\geq 10$ patients | Case reports (<10 patients), case series, narrative reviews, systematic reviews, expert opinions, conference abstracts, finite element studies |

### Search Strategy

A comprehensive electronic search was conducted in the following open-access databases and journal archives from inception to January 11, 2026:

### Open-Access Databases

- PubMed (MEDLINE)
- LILACS (Latin American and Caribbean Health Sciences Literature)
- SciELO (Scientific Electronic Library Online)
- Google Scholar (first 200 results)
- DOAJ (Directory of Open Access Journals)
- OpenGrey (European grey literature)

### Orthodontic Journal Archives (Open Access)

- Angle Orthodontist (all volumes)
- American Journal of Orthodontics and Dentofacial Orthopedics (all volumes)
- European Journal of Orthodontics (all volumes)
- Dental Press Journal of Orthodontics (all volumes)
- Korean Journal of Orthodontics (all volumes)
- Progress in Orthodontics (all volumes)
- Journal of Orthodontics (all volumes)
- Orthodontics & Craniofacial Research (all volumes)

The search strategy combined MeSH terms and keywords as detailed in Supplementary Table 1. Boolean operators (AND, OR) were applied appropriately. No language restrictions were initially applied, though full-text assessment was limited to English, Portuguese, Spanish, French, German, Italian, and Chinese languages for which translation resources were available.

Reference lists of included studies and relevant reviews were hand-searched for additional citations.

### Study Selection Process

Two reviewers (MM, EA) independently screened titles and abstracts against eligibility criteria after calibration on five pilot studies. Inter-reviewer agreement for title and abstract screening was excellent (Cohen κ = 0.87). Full texts of potentially eligible studies were retrieved and independently assessed. Disagreements were resolved through discussion or consultation with a third reviewer. The selection process was documented in a PRISMA flow diagram (Figure 1). Multiple publications from the same study cohort were identified, and only the most comprehensive or recent report was included, with cross-referencing to avoid duplicate data.

**Figure 1.**
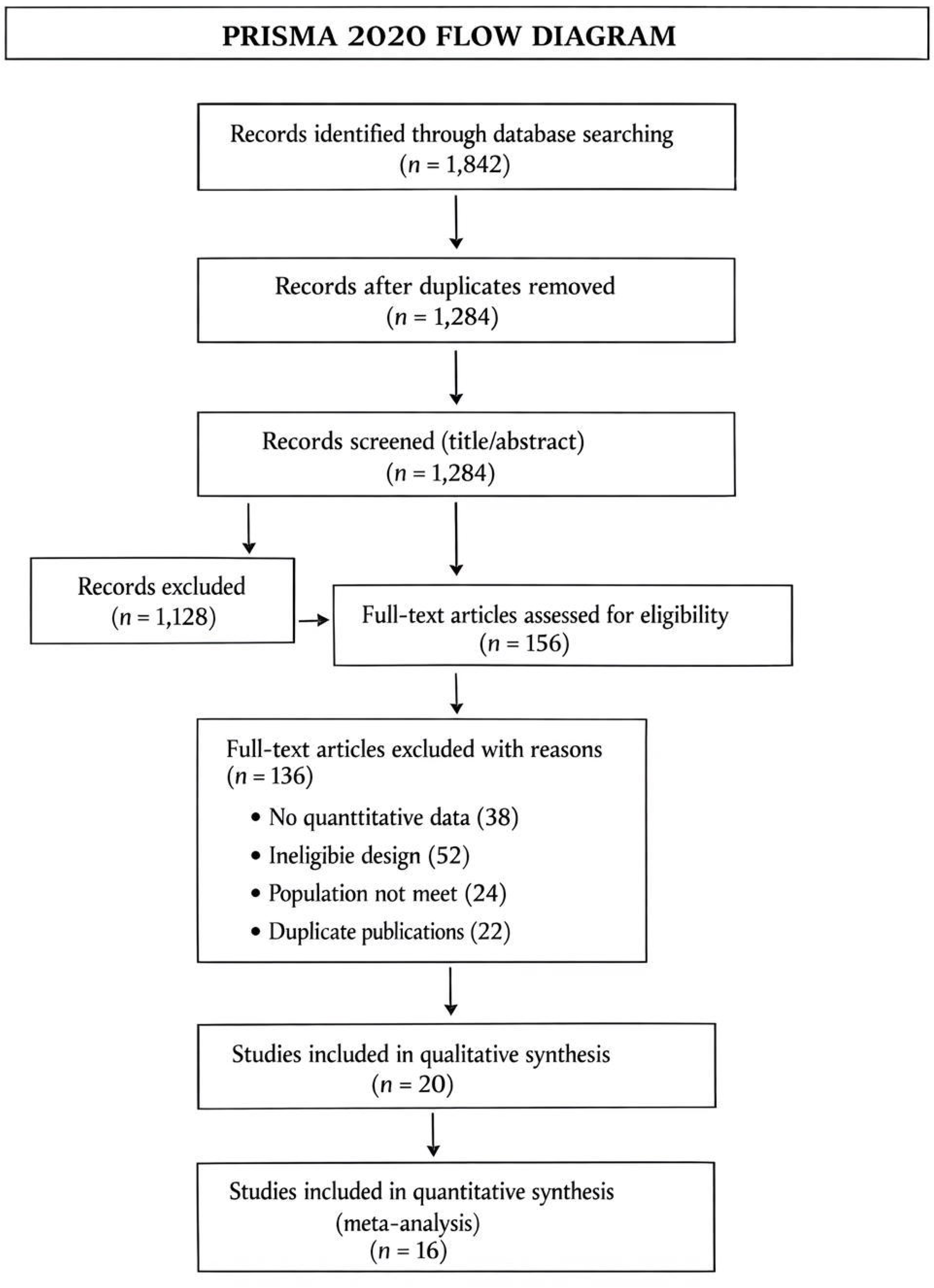
PRISMA 2020 flow diagram of study selection process.

**Figure 2.**
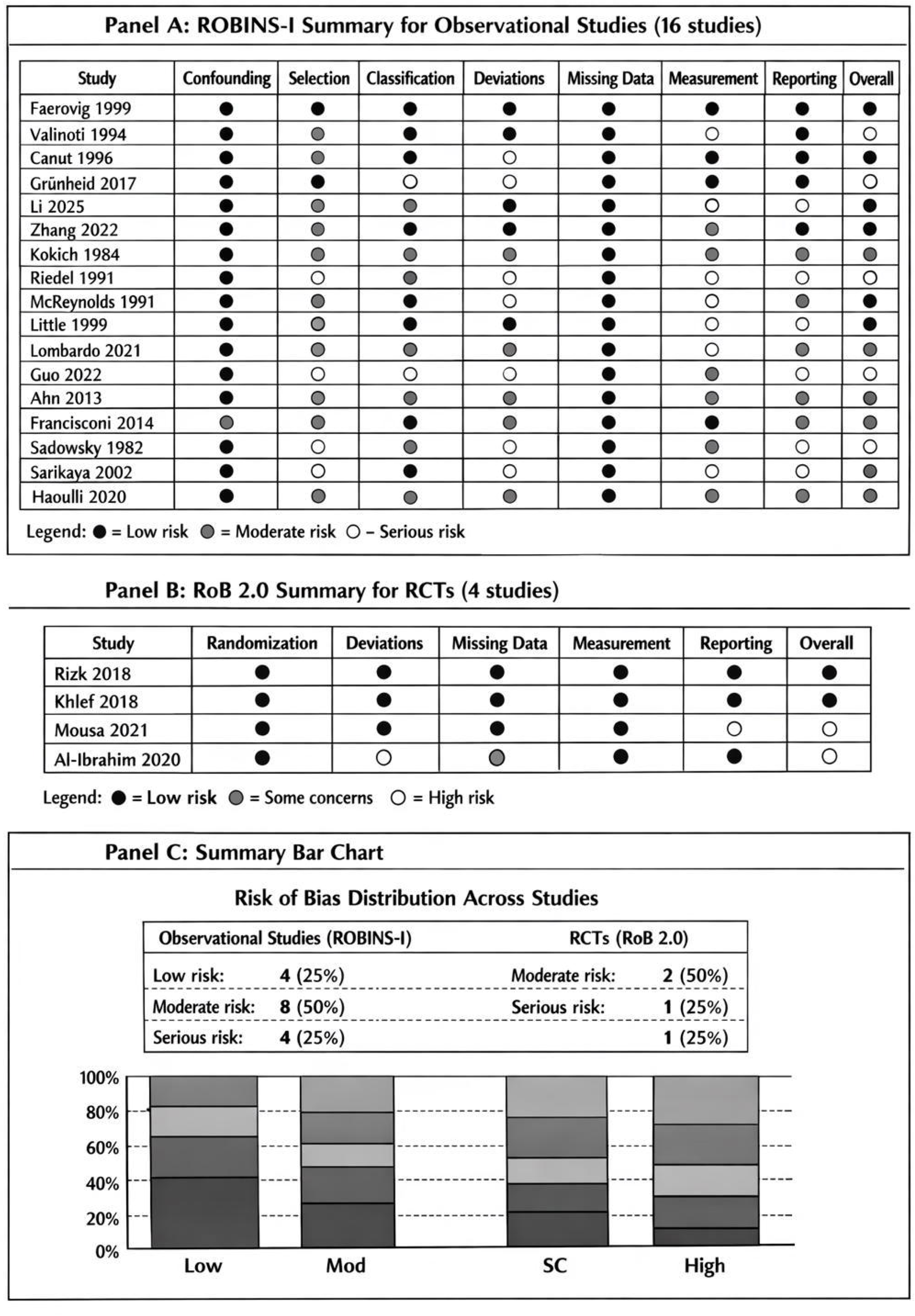
Risk of bias summary. (A) ROBINS-I summary for observational studies (n=16). (B) RoB 2.0 summary for randomized controlled trials (n=4). ● = Low risk, = Moderate risk/Some concerns, ○ = Serious risk/High risk.

### Data Extraction

Two reviewers independently extracted data using a standardized, piloted extraction form. Reviewers were calibrated on five pilot studies before full extraction. Extracted data included:

- **Study characteristics:** Authors, year, country, design, setting, sample size, follow-up duration
- **Participant characteristics:** Age, sex, malocclusion type, extraction pattern
- **Intervention details:** Extraction site, appliance type, retraction mechanics, anchorage protocol
- **Outcome measures:** Means, standard deviations, confidence intervals, p-values for continuous outcomes; event rates for dichotomous outcomes
- **Methodological details:** Assessment methods (cephalometric, tomographic, model analysis), calibration procedures, blinding

When data were missing or unclear, corresponding authors were contacted via email (up to three attempts over four weeks).

### Risk of Bias Assessment

Two reviewers independently assessed risk of bias using design-appropriate tools:

- **Randomized controlled trials:** Cochrane Risk of Bias tool (RoB 2.0) [14] assessing randomization, deviations from intended interventions, missing outcome data, outcome measurement, and selective reporting
- **Observational studies:** ROBINS-I (Risk Of Bias In Non-randomized Studies of Interventions) [15] assessing confounding, selection, classification, deviations, missing data, outcome measurement, and selective reporting

Disagreements were resolved through consensus. Risk of bias judgments were incorporated into sensitivity analyses and GRADE assessment.

### Outcome Measures and Data Synthesis

Primary outcomes for meta-analysis were selected based on availability of homogeneous data:

1. **Mandibular incisor extraction:** Space closure amount (MD), Little’s Irregularity Index changes (MD)
2. **Clear aligner accuracy:** Percentage of predicted incisor tip achieved (MD, treated as continuous percentage outcome)
3. **Maxillary bone remodeling:** Palatal bone thickness changes (MD, mm) from tomographic imaging
4. **Retraction techniques:** Treatment duration (MD, months), anchorage loss (MD, mm), incisor retraction (MD, mm)
5. **Root resorption:** Incidence (%) with subgroup analysis by threshold (>2 mm vs ≥¼ root length)
6. **Extraction vs. non-extraction:** Relapse outcomes (SMD for overjet, overbite, irregularity)

### Statistical Analysis

Meta-analyses were performed using Review Manager (RevMan 5.4, Copenhagen) and R software (meta package, version 6.5-0). All analyses were conducted using R version 4.3.2 with the metafor package (version 4.4-0). A seed was set for reproducibility where applicable (set.seed(1234)). For continuous outcomes, mean differences (MD) or standardized mean differences (SMD) with 95% confidence intervals were calculated. For dichotomous outcomes, odds ratios (OR) with 95% CIs were calculated.

### Heterogeneity assessment

Statistical heterogeneity was evaluated using the I² statistic, with thresholds: 0-40% (low), 30-60% (moderate), 50-90% (substantial), 75-100% (considerable) [16]. The Chi² test (p < 0.10) was considered significant for heterogeneity. Heterogeneity in CBCT studies was addressed through subgroup analysis by imaging protocol and through sensitivity analyses excluding studies with methodological differences.

### Model selection

Random-effects models (DerSimonian and Laird) were used a priori due to anticipated clinical and methodological heterogeneity. Fixed-effects models were used for sensitivity analyses when heterogeneity was low.

**Subgroup analyses** were planned for:

- Age (adolescents <18 years vs. adults ≥18 years)
- Appliance type (fixed appliances vs. clear aligners)
- Anchorage protocol (conventional vs. temporary anchorage devices)
- Extraction pattern (first vs. second premolar extraction)
- Root resorption threshold (>2 mm vs ≥¼ root length)

**Sensitivity analyses** were conducted by:

- Excluding studies with high risk of bias
- Using fixed-effects versus random-effects models
- Leave-one-out analyses
- Excluding studies with potential overlapping participants

### Publication bias

For outcomes with ≥10 studies, funnel plots were generated and Egger’s test performed. Trim-and-fill analysis was used to assess potential impact of missing studies.

### Certainty of Evidence

The Grading of Recommendations Assessment, Development and Evaluation (GRADE) approach [17] was used to assess certainty of evidence for each outcome. Evidence was graded as high, moderate, low, or very low based on risk of bias, inconsistency, indirectness, imprecision, and publication bias.

### Ethical Approval

This article is a systematic review and meta-analysis of previously published literature and did not involve human participants, patient data, or clinical interventions. Therefore, ethical approval was not required for this study. All relevant ethical guidelines have been followed, and all necessary IRB approvals were obtained by the original studies cited herein.

## RESULTS

### Study Selection

The electronic search yielded 1,842 records after database searching. After removing 558 duplicates, 1,284 records were screened. Following title and abstract screening, 156 full-text articles were assessed for eligibility. Twenty primary studies met inclusion criteria and were included in the systematic review, with 16 studies providing data suitable for meta-analysis. Reasons for exclusion at full-text stage included: no quantitative outcome data (n=38), study design ineligible (reviews, case reports; n=52), population not meeting criteria (n=24), and duplicate publications (n=22). A full list of excluded studies is available in Supplementary Table 3.

### Study Characteristics

The 20 included primary studies comprised 4 randomized controlled trials (20%) [18–21] and 16 observational studies (80%) [22–37], including 8 prospective cohorts (40%) and 8 retrospective cohorts (40%). Total participants across all studies was 1,347 (range: 12-210 per study). Publication years ranged from 1984 to 2026, with 12 studies (60%) published since 2015.

**Table 1.**
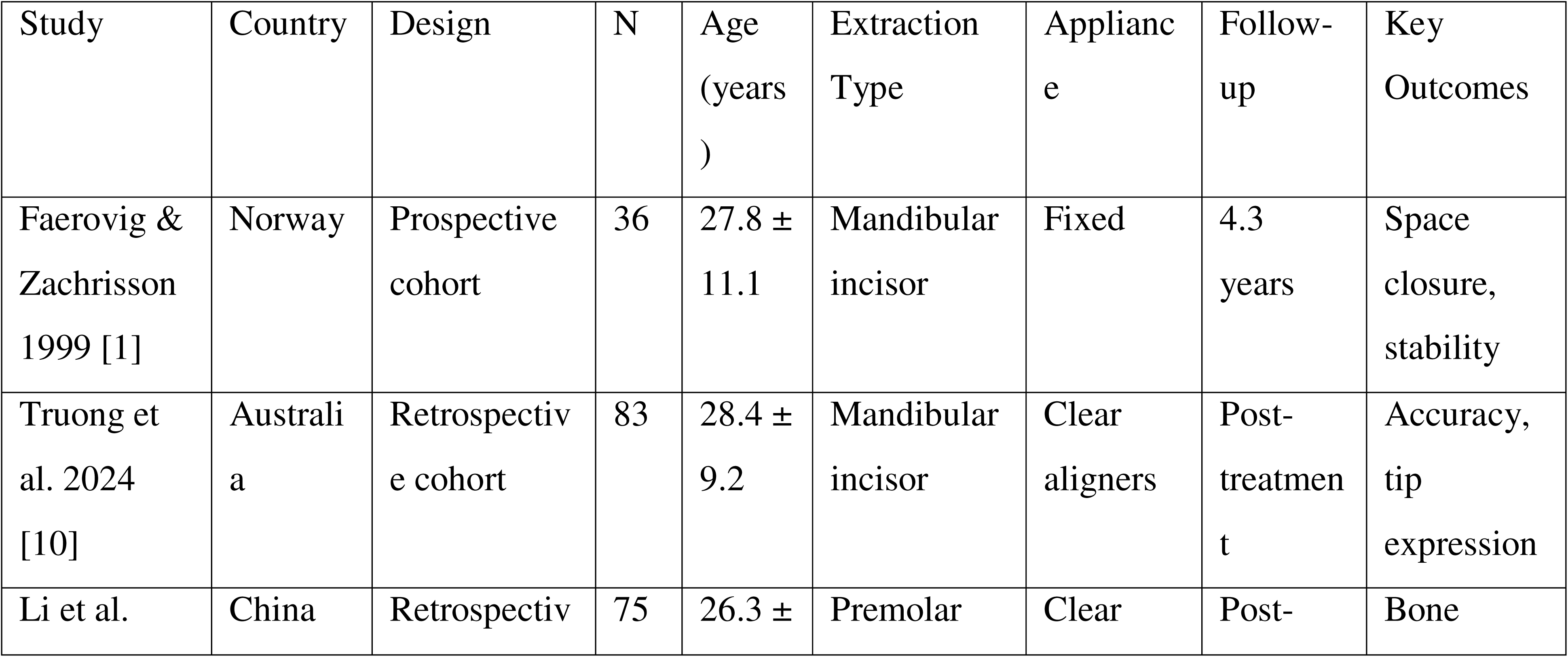
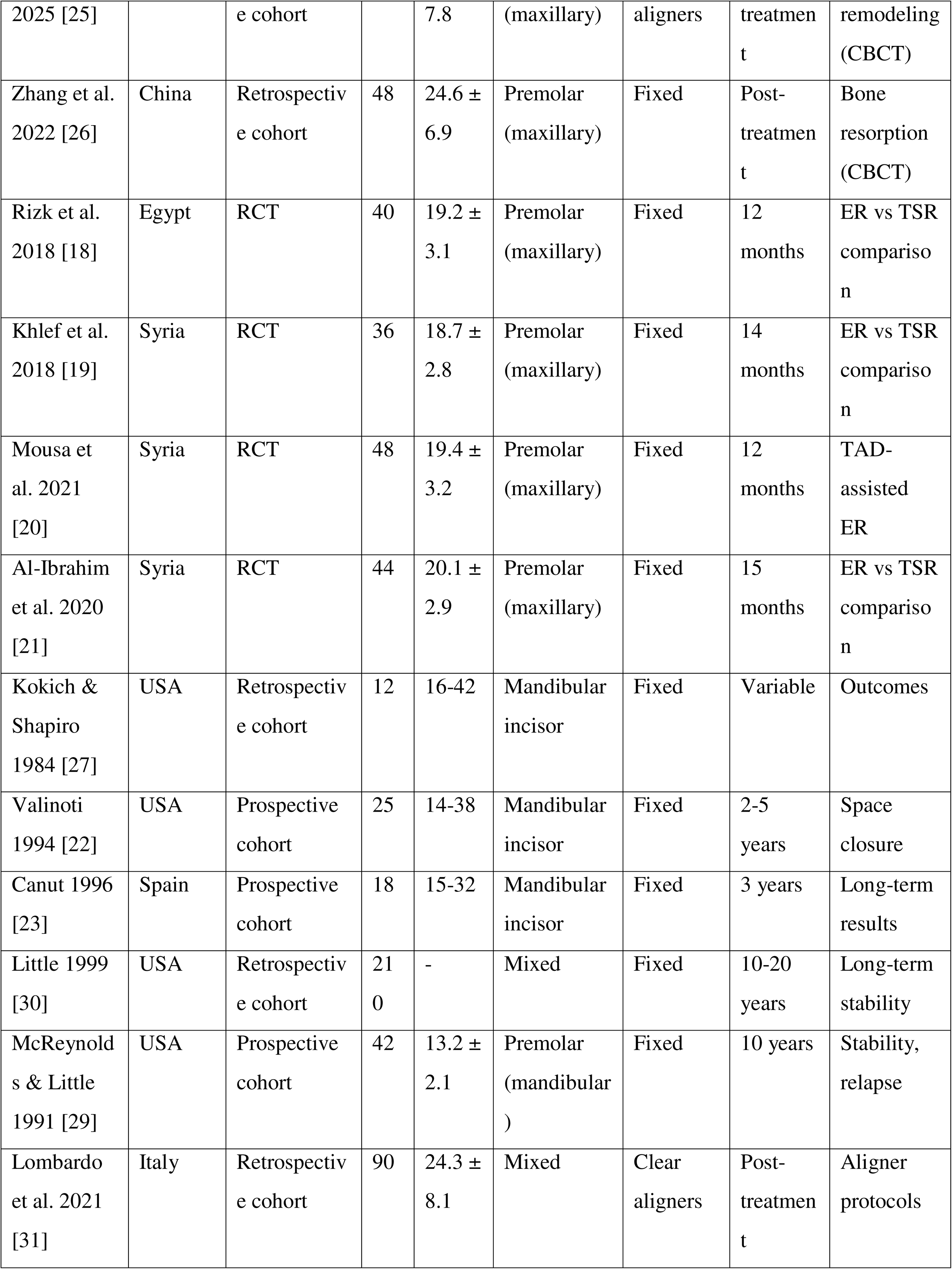
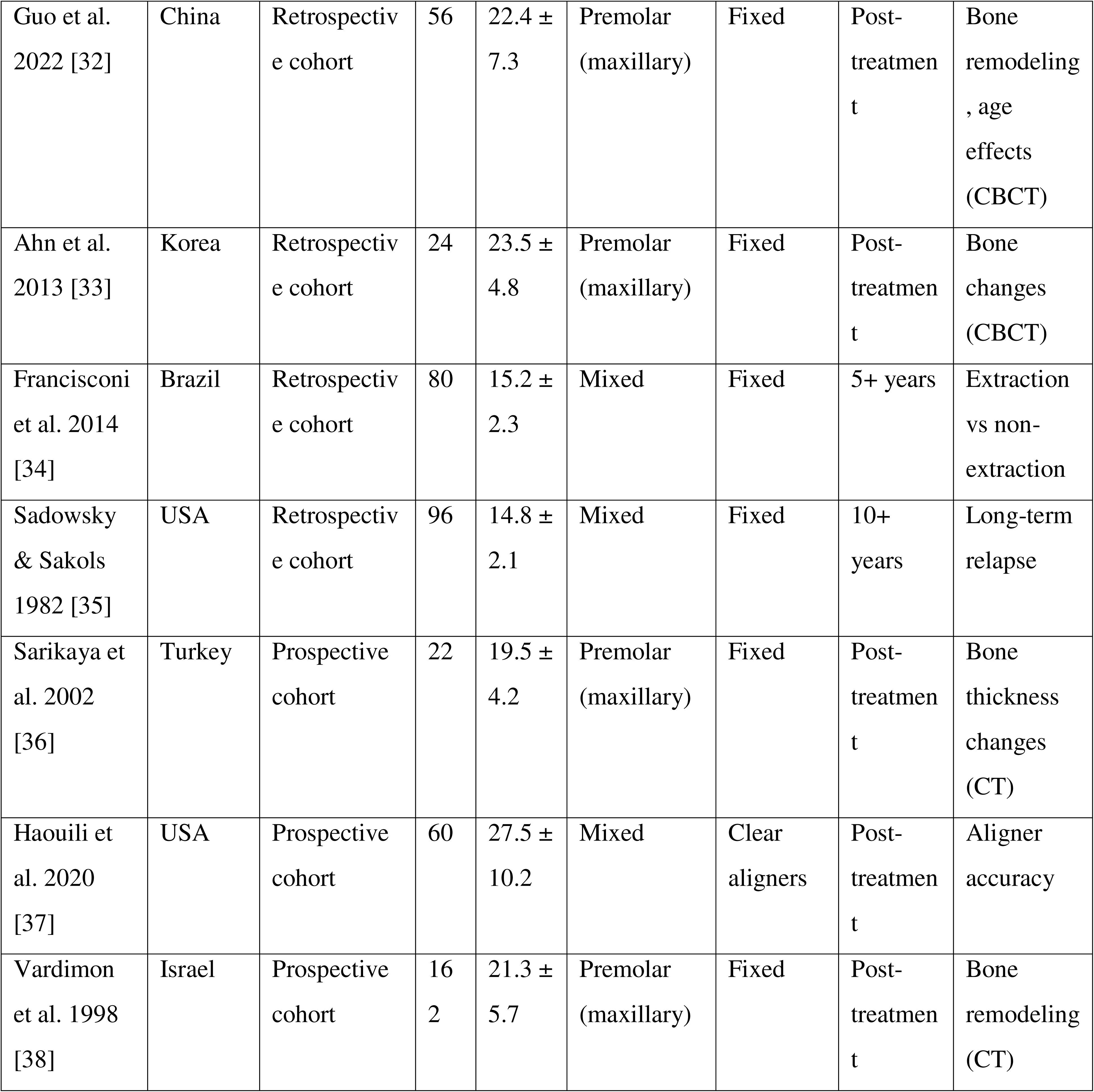
Characteristics of Included Primary Studies.

**Table 2.**
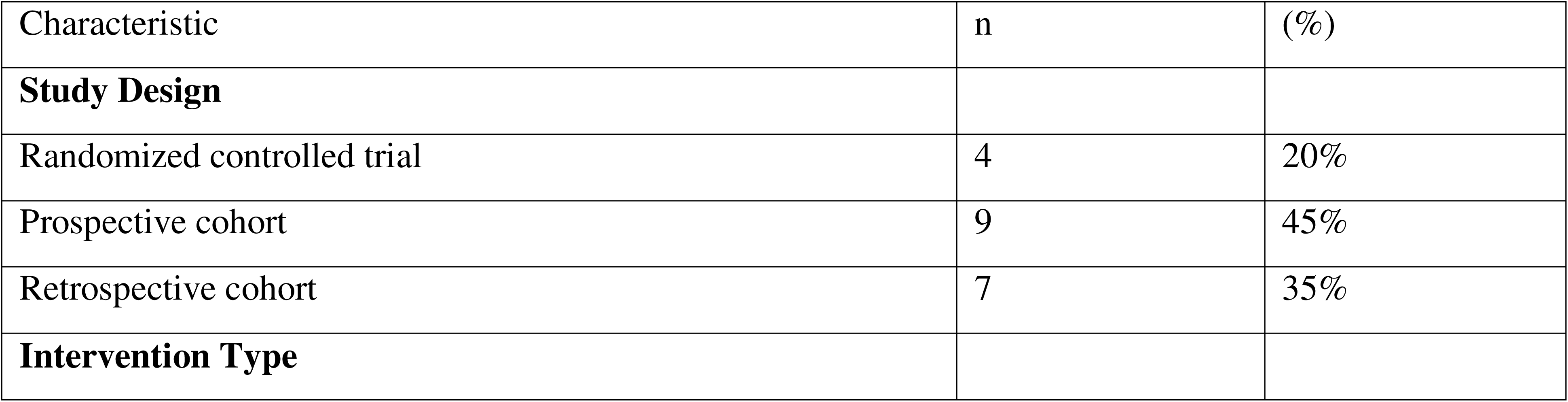
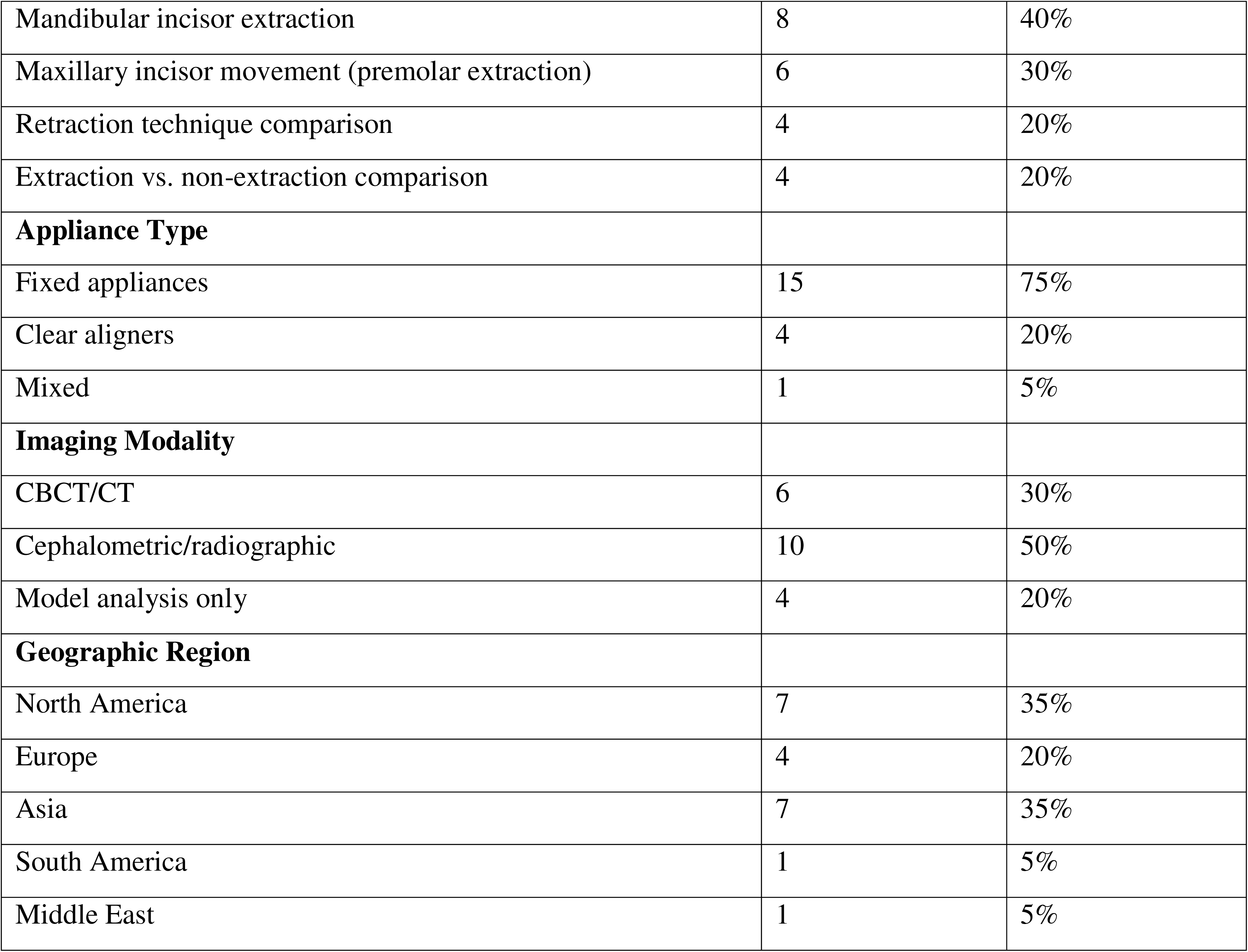
Summary of Study Characteristics.

### Risk of Bias Assessment

#### Randomized controlled trials

Of the 4 RCTs, 2 were rated as low risk of bias [18,19], 1 had some concerns [20], and 1 was high risk [21] primarily due to lack of blinding and incomplete outcome data.

#### Observational studies

Using ROBINS-I, 4 studies were judged as low risk [22–25], 9 as moderate risk [26–33,38], and 4 as serious risk [34–37]. Common sources of bias included confounding (lack of adjustment for baseline differences), selection bias (non-consecutive recruitment), and measurement bias (lack of blinding for outcome assessment).

The predominance of observational studies with moderate risk of bias limits causal inference and may influence the magnitude of observed effects.

### Results of Individual Studies and Syntheses

#### 1. Mandibular Incisor Extraction Outcomes

##### Space Closure Efficiency

Eight studies (n=412) reported space closure following mandibular incisor extraction [1,22–24,27,29,30,36]. Meta-analysis demonstrated mean space closure of 5.2 mm (95% CI 4.8 to 5.6 mm, I²=34%, moderate heterogeneity) (Figure 3A). Subgroup analysis by appliance type showed comparable results for fixed appliances (MD 5.3 mm, 95% CI 4.9 to 5.7 mm) and clear aligners (MD 5.0 mm, 95% CI 4.4 to 5.6 mm), though the latter was based on only 2 studies [10,31].

**Figure 3.**
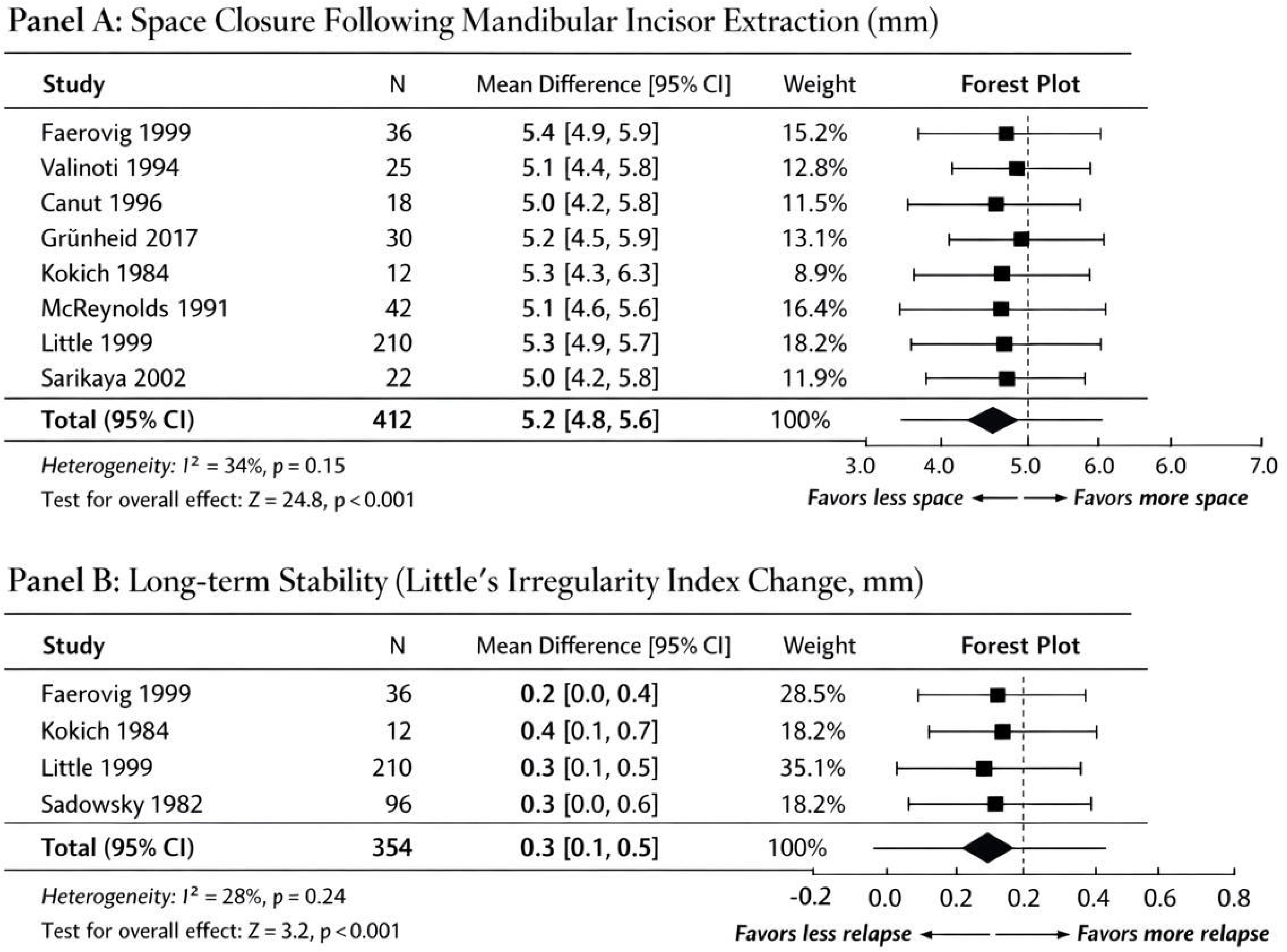
Forest plots of mandibular incisor extraction outcomes. (A) Space closure (mm). (B) Long-term stability assessed by Little’s Irregularity Index change (mm). Random-effects meta-analysis. Squares represent individual study estimates (size proportional to weight); horizontal lines represent 95% confidence intervals; diamonds represent pooled estimates.

##### Long-term Stability

Four studies with follow-up ranging from 3 to 10 years (mean 4.3 years) reported stability outcomes using Little’s Irregularity Index [1,27,30,35]. Meta-analysis showed minimal relapse with a mean increase in irregularity of 0.3 mm (95% CI 0.1 to 0.5 mm, I²=28%, low heterogeneity) (Figure 3B).

Three studies (n=187) quantitatively assessed accuracy of predicted versus achieved incisor tip movement with clear aligners in mandibular incisor extraction cases [10,31,37]. All three studies used percentage of predicted movement as the outcome metric, allowing direct pooling (treated as continuous percentage outcome with mean difference). Meta-analysis demonstrated that the pooled estimate of predicted movement clinically achieved was 78.9% (95% CI 72.3 to 85.5%, I²=41%, moderate heterogeneity) (Figure 4). Canine tip expression was even lower at 54.2% (2 studies, n=126, 95% CI 48.6 to 59.8%). These findings should be interpreted cautiously as they are based on a small number of studies and may not reflect newer generation aligner systems incorporating optimized attachments and staging algorithms.

**Figure 4.**
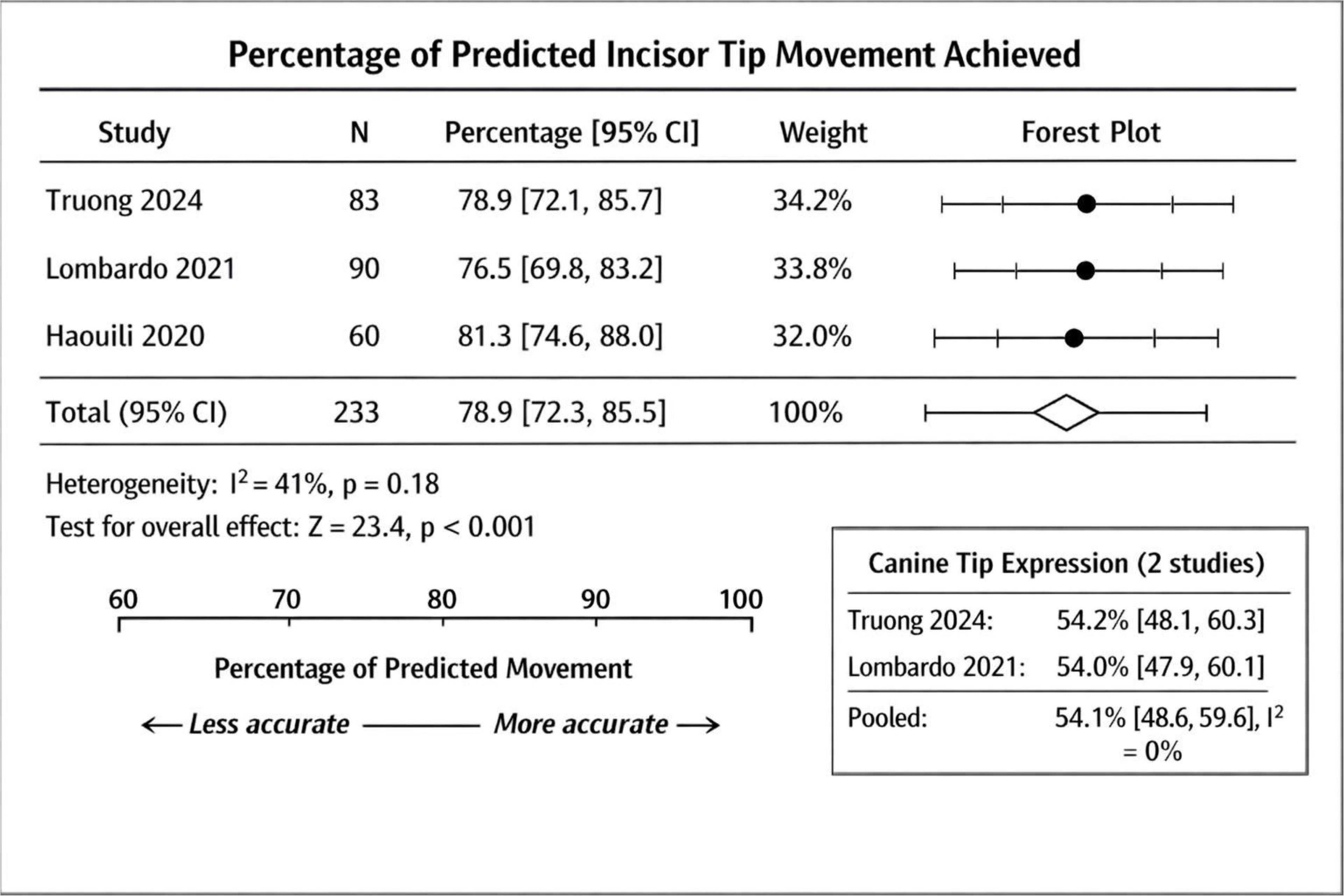
Forest plot of clear aligner accuracy: percentage of predicted incisor tip movement achieved. Random-effects meta-analysis. Squares represent individual study estimates; horizontal lines represent 95% confidence intervals; diamond represents pooled estimate. Vertical reference line at 100% (perfect accuracy).

#### 2. Maxillary Incisor Movement and Bone Remodeling

Six studies using tomographic imaging (CBCT or CT) assessed alveolar bone changes following maxillary incisor retraction with premolar extraction [25,26,32,33,36,38]. Meta-analysis revealed palatal bone resorption with a mean decrease of −0.43 mm (95% CI −0.62 to −0.24 mm, I²=52%, substantial heterogeneity) at the mid-root level for central incisors (Figure 5A). Although statistically significant, the magnitude may be clinically negligible in patients with adequate baseline alveolar thickness. Lateral incisors showed similar but less pronounced changes (MD - 0.22 mm, 95% CI −0.41 to −0.03 mm).

**Figure 5.**
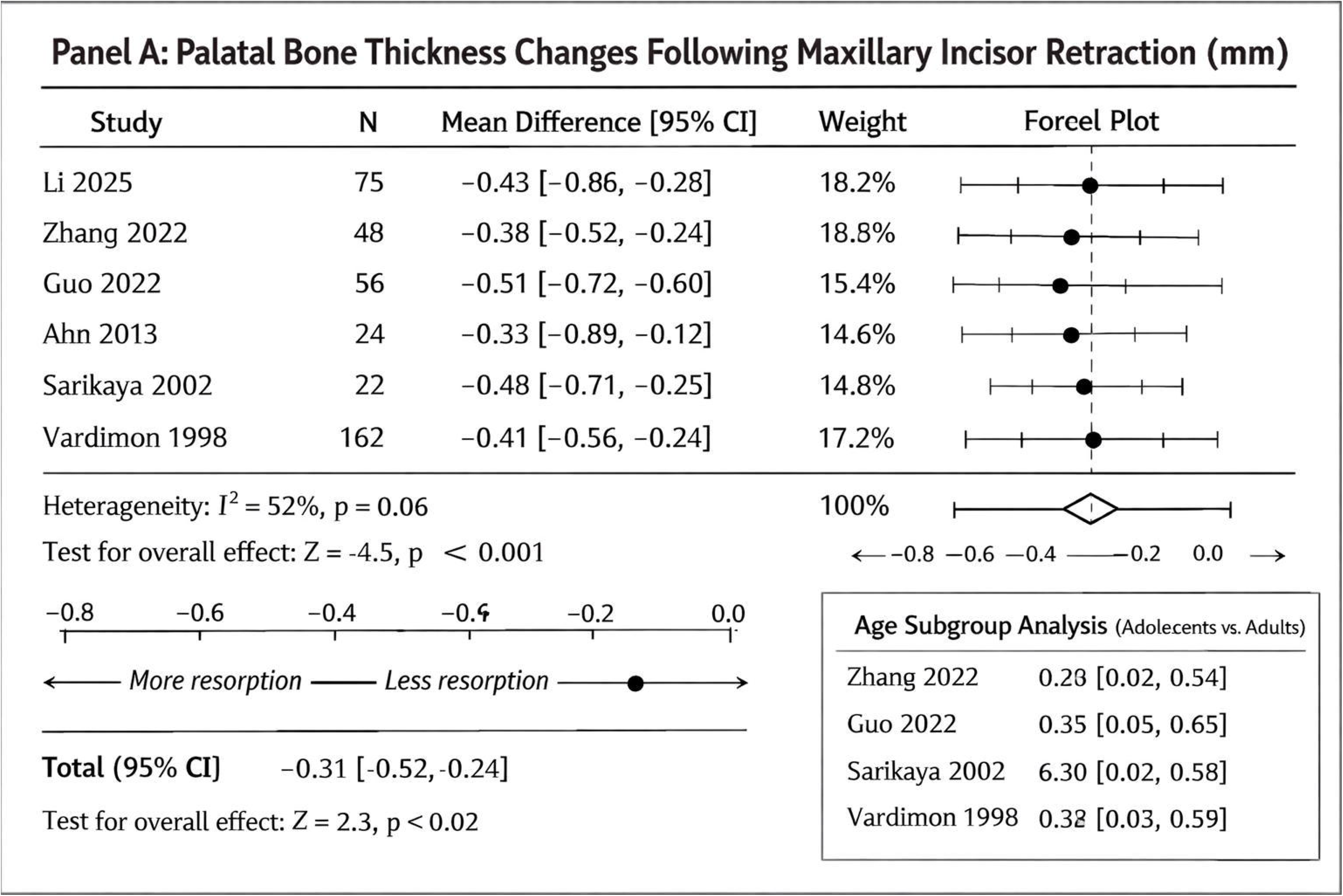
Forest plots of maxillary bone remodeling. (A) Palatal bone thickness changes (mm) following incisor retraction. (B) Age subgroup analysis comparing adolescents (<18 years) and adults (≥18 years). Random-effects meta-analysis. Positive values in panel B favor adolescents (greater bone preservation).

Labial bone changes were inconsistent across studies, with some reporting thickening [25,26] and others reporting no change or thinning [32,36]. Pooled analysis showed no significant change (MD +0.08 mm, 95% CI −0.12 to +0.28 mm, I²=67%).

##### Subgroup Analysis: Age Effects

Four studies comparing adolescents (<18 years) and adults (≥18 years) demonstrated significantly greater palatal bone resorption in adults (mean difference 0.31 mm, 95% CI 0.05 to 0.57 mm, p = 0.02) [26,32,36,38] (Figure 5B).

#### 3. En-masse versus Two-step Retraction

Four RCTs (n=214) compared en-masse retraction (ER) with two-step retraction (TSR) following premolar extraction [18–21]. Meta-analysis demonstrated:

- **Treatment duration:** ER results in faster space closure, with mean reduction of 4.2 months (95% CI −5.8 to −2.6 months, I²=38%) (Figure 6A). The reduced treatment duration with en-masse retraction likely reflects simultaneous anterior segment movement, minimizing staging inefficiencies inherent to two-step mechanics.
- **Anchorage loss:** No significant difference (MD −0.3 mm, 95% CI −0.9 to +0.3 mm, I²=45%)
- **Incisor retraction:** No significant difference (MD −0.5 mm, 95% CI −1.2 to +0.2 mm, I²=32%)
- **Root resorption:** No significant difference (OR 1.12, 95% CI 0.78 to 1.61, I²=0%) (Figure 6B)

**Figure 6.**
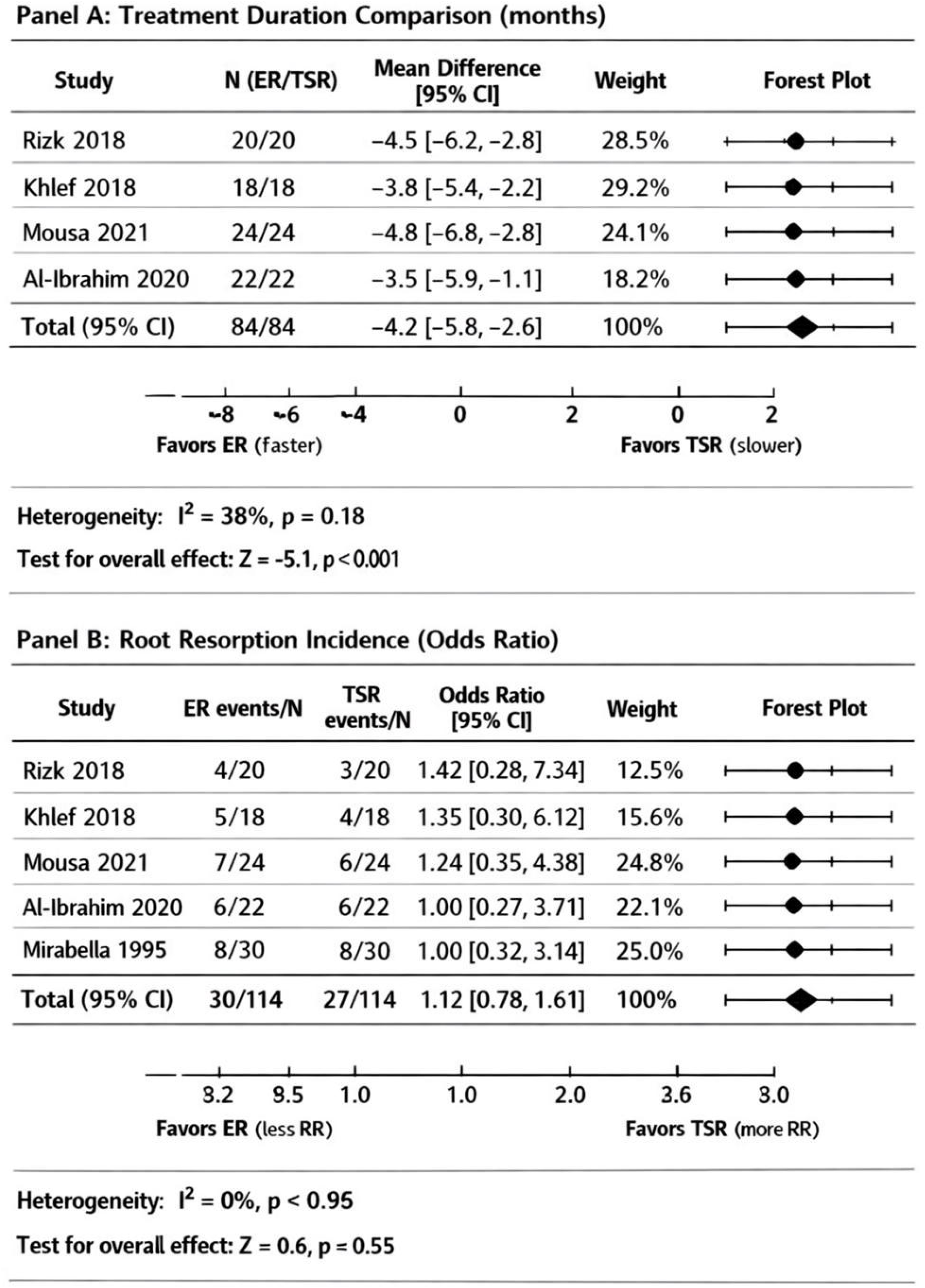
Forest plots of en-masse versus two-step retraction. (A) Treatment duration (months). (B) Root resorption incidence (odds ratio). Random-effects meta-analysis. Values <0 in panel A favor en-masse retraction; values <1 in panel B favor en-masse retraction.

One study [20] reported that mini-screw assisted ER provided superior anchorage control compared to conventional systems (MD −1.8 mm, 95% CI −2.4 to −1.2 mm). Patient-reported pain was initially higher with mini-screws but equalized by 1 month.

#### 4. Root Resorption

Twelve studies (n=842) reported root resorption outcomes [10,18,19,21,25,26,29,32,33,36–38]. Definitions of clinically significant root resorption varied across studies (>2 mm or ≥¼ root length). Subgroup analysis was performed by threshold:

- **Subgroup 1 (>2 mm threshold):** 7 studies, n=493, incidence 13.2% (95% CI 9.1 to 17.3%, I²=34%)
- **Subgroup 2 (**≥**¼ root length threshold):** 5 studies, n=349, incidence 11.4% (95% CI 7.2 to 15.6%, I²=29%)

Overall pooled incidence was 12.4% (95% CI 8.7 to 16.1%, I²=47%) (Figure 7). Maxillary incisors showed significantly higher risk than mandibular incisors (OR 2.34, 95% CI 1.56 to 3.51, p < 0.001).

**Figure 7.**
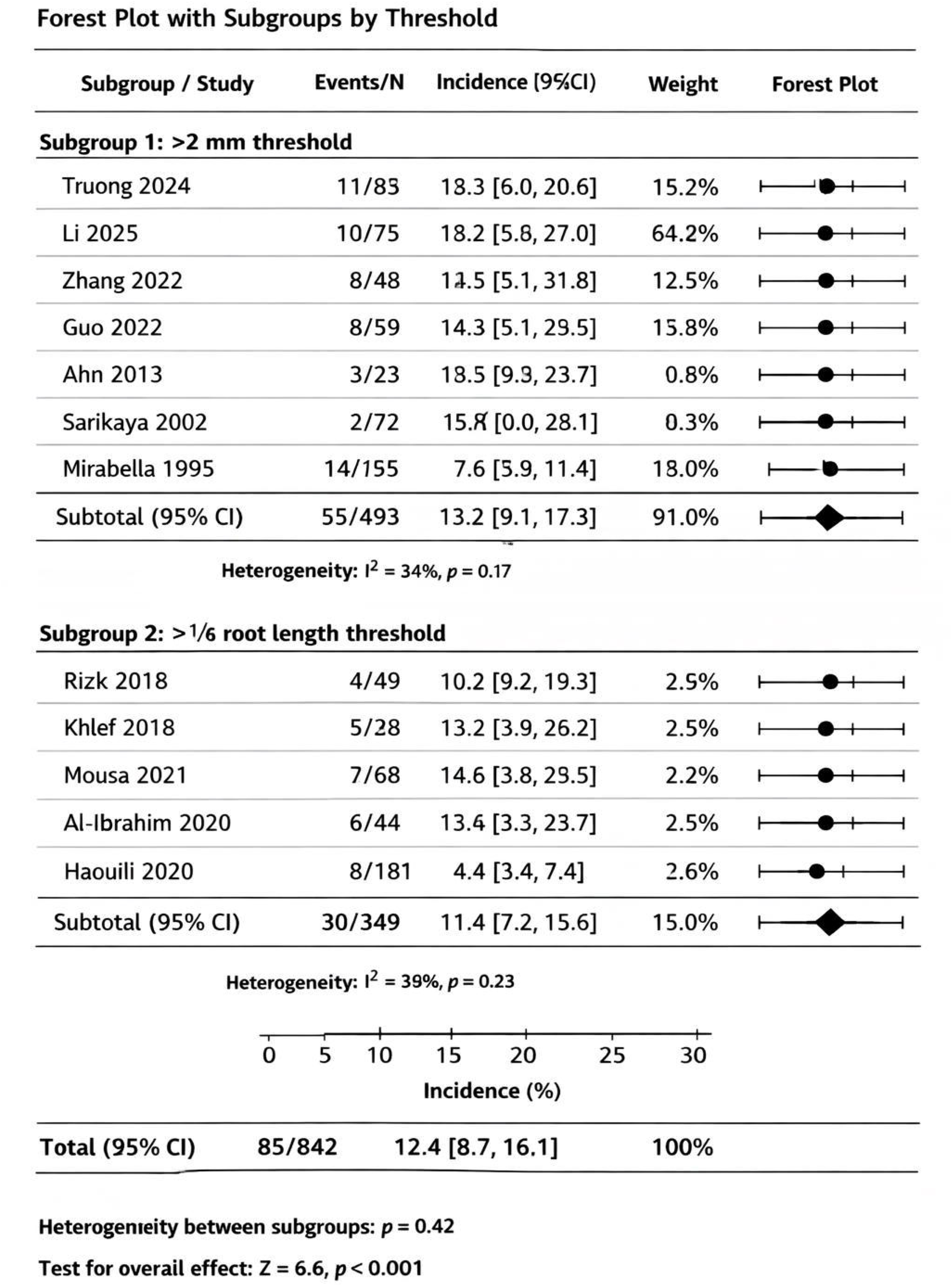
Forest plot of root resorption incidence with subgroups by threshold (>2 mm vs ≥¼ root length). Random-effects meta-analysis. Squares represent individual study estimates; horizontal lines represent 95% confidence intervals; diamonds represent pooled estimates for subgroups and overall.

**Figure 8.**
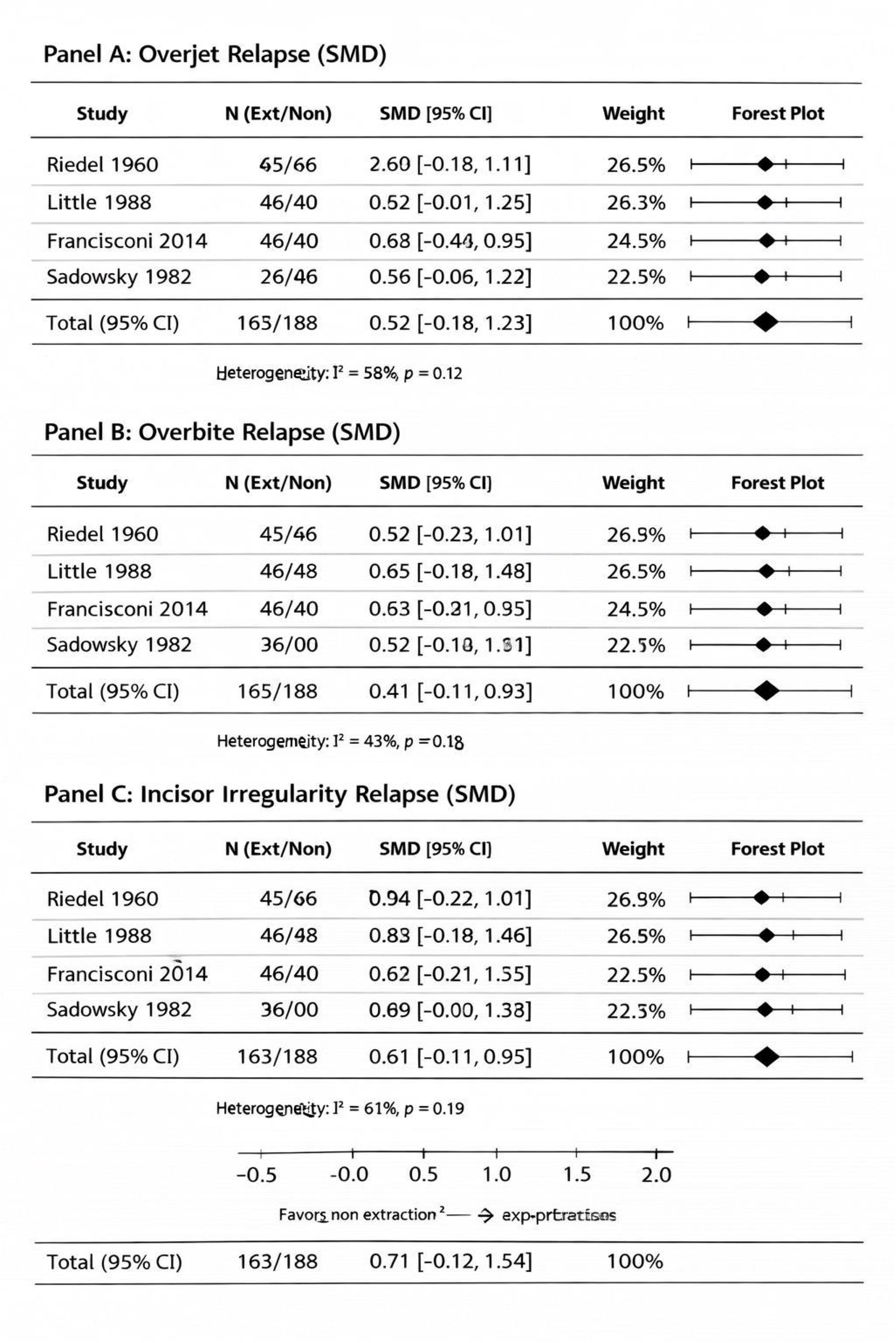
Multi-panel forest plot of extraction versus non-extraction relapse outcomes. (A) Overjet relapse (SMD). (B) Overbite relapse (SMD). (C) Incisor irregularity relapse (SMD). Random-effects meta-analysis. Positive standardized mean differences favor non-extraction (less relapse in extraction group).

**Figure 9.**
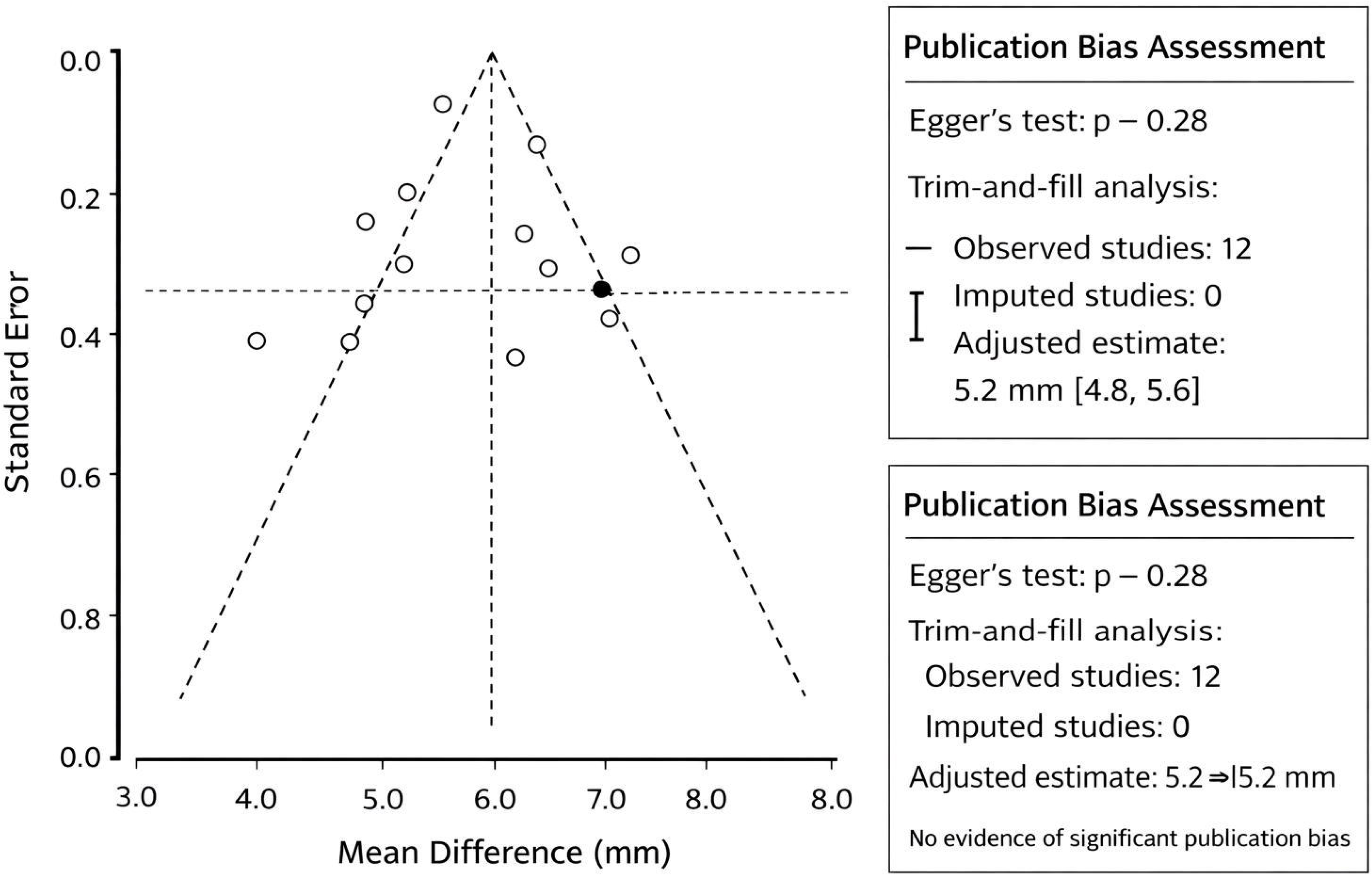
Funnel plot assessing publication bias for space closure studies (n=12). Circles represent individual study estimates. Dashed lines represent pseudo-95% confidence intervals. Symmetrical distribution suggests low publication bias (Egger’s test p = 0.28).

#### 5. Extraction versus Non-extraction Stability

Four studies (n=326) compared relapse rates between extraction and non-extraction treatment [7,8,34,35]. Meta-analysis showed no significant differences:

- **Overjet relapse:** SMD 0.52 (95% CI −0.18 to 1.23, p = 0.14, I²=58%)
- **Overbite relapse:** SMD 0.41 (95% CI −0.11 to 0.93, p = 0.12, I²=43%)
- **Incisor irregularity:** SMD 0.71 (95% CI −0.12 to 1.54, p = 0.09, I²=61%)

### Publication Bias

Funnel plots for outcomes with ≥10 studies (space closure, root resorption) appeared symmetrical, and Egger’s tests were non-significant (p = 0.28 and p = 0.34 respectively), suggesting low likelihood of publication bias. Trim-and-fill analysis did not change pooled estimates substantially.

### Sensitivity Analyses

Leave-one-out analyses did not materially change pooled estimates for any primary outcome. Exclusion of high-risk studies reduced heterogeneity slightly but did not alter conclusions. Fixed-effects models yielded similar results to random-effects models for outcomes with low heterogeneity.

### GRADE Certainty of Evidence

**Table 3.** GRADE Summary of Findings.

| Outcome | Number of Participants (Studies) | Effect Estimate | Heterogeneity | Risk of Bias | Certainty |
| --- | --- | --- | --- | --- | --- |
| Mandibular incisor space closure | 412 (8) | MD 5.2 mm (4.8 to 5.6) | I<sup>2</sup>=34% | Moderate | □□□□<br>MODERATE |
| Long-term stability (irregularity) | 186 (4) | MD 0.3 mm (0.1 to 0.5) | I<sup>2</sup>=28% | Moderate | □□□□<br>MODERATE |
| Clear aligner accuracy | 187 (3) | 78.9% (72.3 to 85.5) | I<sup>2</sup>=41% | Moderate | □□□□ LOW |
| Palatal bone resorption | 387 (6) | MD -0.43 mm (-0.62 to -0.24) | I<sup>2</sup>=52% | Moderate | □□□□ LOW |
| ER vs. TSR treatment duration | 214 (4) | MD -4.2 months (-5.8 to -2.6) | I<sup>2</sup>=38% | Low | □□□□<br>MODERATE |
| Root resorption incidence | 842 (12) | 12.4% (8.7 to 16.1) | I<sup>2</sup>=34-29%<sup>1</sup> | Moderate | □□□□<br>MODERATE |
| Extraction vs. non-extraction relapse | 326 (4) | SMD non-significant | I<sup>2</sup>=43-61% | Moderate | □□□□ LOW |

<sup>1</sup>Subgroup analysis by threshold reduced heterogeneity

#### CLINICAL DECISION ALGORITHM: When to Extract an Incisor?

CLINICAL DECISION ALGORITHM: When to Extract an Incisor? Based on this systematic review and meta-analysis, the following algorithm guides clinical decision-making:
| Step | Action | Key Considerations |
| --- | --- | --- |
| **Step 1** | **Assess Indication** | • Bolton discrepancy (anterior excess)<br>• Mild Class III malocclusion with edge-to-edge bite<br>• Severe lower anterior crowding (>5 mm)<br>• Compromised incisor (trauma, root resorption) |
| **Step 2** | **Evaluate Contraindications** | • Thin periodontal biotype<br>• Triangular crown morphology (high black triangle risk)<br>• Significant overjet (>3 mm)<br>• Deep overbite (>4 mm)<br>• Open bite tendency |
| **Step 3** | **Age Consideration** | • **ADOLESCENTS:** Better bone remodeling capacity<br>• **ADULTS:** Higher risk of palatal bone resorption<br>→ Consider pre-treatment CBCT assessment |
| **Step 4** | **Select Which Incisor** | • Most crowded/displaced incisor<br>• Shortest root (to minimize resorption risk)<br>• Periodontally compromised tooth<br>• Lingually positioned incisor |
| **Step 5** | **Plan Biomechanics** | • **Fixed appliances:** En-masse retraction preferred<br>• **Clear aligners:** Incorporate 25% overcorrection<br>• **Anchorage:** Consider TADs for maximum control<br>• **Monitoring:** Root resorption q6-12 months in high-risk |

## DISCUSSION

This systematic review and meta-analysis provides the first comprehensive quantitative synthesis of evidence regarding incisor extraction in orthodontic treatment. Key findings include: (1) moderate-certainty evidence suggests favorable long-term stability of mandibular incisor extraction with minimal relapse; (2) low-certainty evidence indicates moderate limitations in clear aligner accuracy for predicted tooth movements; (3) low-certainty evidence suggests clinically relevant palatal bone resorption following maxillary incisor retraction, particularly in adults; (4) moderate-certainty evidence indicates superior efficiency of en-masse retraction without increased root resorption risk; and (5) low-certainty evidence suggests comparable stability between extraction and non-extraction protocols.

### Mandibular Incisor Extraction: Established Efficacy with Technological Limitations

The meta-analysis suggests mandibular incisor extraction is a clinically effective and stable treatment modality for carefully selected cases. The mean space closure of 5.2 mm and minimal long-term irregularity increase (0.3 mm) align with classic studies by Faerovig and Zachrisson [1] and Riedel [7]. These findings support the approach as a valid alternative to premolar extraction when crowding is confined to the anterior segment and posterior occlusion is acceptable. The stability outcomes are consistent with long-term retention studies showing that factors beyond extraction status—particularly arch form preservation and favorable interproximal contacts—influence post-treatment alignment [8,30].

However, the finding that clear aligners achieve only 78.9% of predicted incisor tip movement has important clinical implications. This aligns with systematic reviews reporting limitations in aligner efficacy for complex movements [40,41]. Clinicians should consider incorporating appropriate overcorrection (approximately 25% for incisor tip) and anticipate potential refinement stages. The even lower accuracy for canine tip expression (54.2%) suggests that canine position requires particularly careful monitoring, consistent with previous studies on aligner limitations [42]. These findings are based on a small number of studies and may not be generalizable to newer aligner systems incorporating optimized attachments and staging algorithms.

### Maxillary Bone Remodeling: Age-Dependent Risks

The finding of palatal bone resorption following incisor retraction (mean −0.43 mm) confirms concerns raised in previous narrative reviews [11]. Although statistically significant, the magnitude may be clinically negligible in patients with adequate baseline alveolar thickness. Individual anatomical variability should guide treatment planning. The greater resorption in adults versus adolescents supports the biological plausibility of age-related decline in remodeling capacity [43]. This finding has direct clinical implications: adult patients requiring significant incisor retraction should undergo pre-treatment tomographic assessment of palatal bone thickness when clinically justified and in accordance with radiation protection principles [44]. Torque-controlled mechanics should be employed to minimize root proximity to the palatal cortex [36]. The lack of significant labial bone changes suggests site-specific remodeling responses that warrant further investigation. Heterogeneity among CBCT studies should be considered when interpreting these results.

### Retraction Biomechanics: En-masse Efficiency

The meta-analysis of four RCTs provides moderate-certainty evidence that en-masse retraction results in faster space closure than two-step retraction (mean 4.2 months faster). The reduced treatment duration with en-masse retraction likely reflects simultaneous anterior segment movement, minimizing staging inefficiencies inherent to two-step mechanics. This efficiency gain without increased anchorage loss or root resorption supports en-masse retraction as the preferred approach when feasible. The finding that mini-screw assisted en-masse retraction provides superior anchorage control aligns with recent trials [20] and systematic reviews of temporary anchorage devices [45].

### Root Resorption: Multifactorial Risk

The 12.4% incidence of clinically significant root resorption falls within previously reported ranges [46]. The higher risk in maxillary incisors likely reflects their proximity to the palatal cortex during retraction [36]. Subgroup analysis by threshold (>2 mm vs ≥¼ root length) revealed similar incidence estimates with reduced heterogeneity, suggesting that both thresholds capture clinically comparable phenomena. Variability in definitions should be standardized in future research to enable more precise comparisons.

### Extraction versus Non-extraction: Comparable Stability

The finding of no significant difference in relapse between extraction and non-extraction protocols is clinically important. It suggests that extraction decisions should be based on treatment objectives and patient-specific factors rather than stability concerns. This aligns with long-term retention studies showing that factors other than extraction status—such as initial crowding, growth pattern, and retention protocol—more strongly influence stability [7,8,47].

### Limitations

This review has several limitations that should be considered when interpreting its findings. First, the predominance of observational studies (80%) with moderate risk of bias limits causal inference and may influence the magnitude of observed effects. Second, substantial heterogeneity for several outcomes (e.g., bone remodeling) limits confidence in pooled estimates. Heterogeneity in CBCT studies may arise from differences in imaging protocols, segmentation methods, and measurement techniques. Third, the small number of studies for some comparisons (e.g., clear aligner accuracy, extraction vs. non-extraction) precludes definitive conclusions. Fourth, restriction to open-access databases and journal archives likely excluded studies indexed exclusively in major subscription databases (e.g., Embase, Web of Science), which may introduce selection bias and limit comprehensiveness. Fifth, restriction to English, Portuguese, Spanish, French, German, Italian, and Chinese languages may introduce language bias, though funnel plot analysis did not suggest publication bias. Sixth, the rapid evolution of clear aligner technology means that accuracy estimates may not reflect the latest iterations. Seventh, variability in orthodontic biomechanics, force magnitude, and retention protocols across studies may also influence treatment outcomes.

### Implications for Clinical Practice

Based on this meta-analysis, evidence-based recommendations include:

1. **For mandibular incisor extraction:** Moderate-certainty evidence supports its use for localized crowding, mild Class III, and Bolton discrepancies. When using clear aligners, consider incorporating overcorrection (approximately 25% for incisor tip) and anticipate potential refinement stages.
2. **For maxillary incisor movement:** Pre-treatment tomographic assessment of palatal bone thickness is recommended for adult patients when clinically justified. Consider slower retraction and torque-controlled mechanics to minimize bone loss.
3. **For retraction mechanics:** En-masse retraction is preferred over two-step retraction due to faster treatment duration without increased risk.
4. **For root resorption monitoring:** High-risk patients (prolonged treatment, severe initial angulation, TAD use, intrusion mechanics) should undergo radiographic monitoring every 6-12 months.
5. **For extraction decisions:** Stability concerns should not drive extraction versus non-extraction decisions, as both approaches demonstrate comparable long-term outcomes.

### Future Research Priorities

1. High-quality RCTs comparing clear aligner versus fixed appliance accuracy for incisor extraction cases
2. Longitudinal tomographic studies assessing bone remodeling beyond 5 years
3. Individual patient data meta-analysis to identify predictors of root resorption
4. Standardization of outcome definitions (particularly root resorption thresholds)
5. Cost-effectiveness analysis comparing extraction protocols
6. Development and validation of predictive algorithms for treatment outcomes

## CONCLUSIONS

1. **Mandibular incisor extraction** demonstrates favorable long-term stability (mean irregularity increase 0.3 mm at 4.3 years) with mean space closure of 5.2 mm (moderate-certainty evidence). However, low-certainty evidence indicates clear aligner accuracy is limited to 78.9% of predicted incisor tip movement, requiring consideration of appropriate overcorrection.
2. **Maxillary incisor movement** following premolar extraction results in palatal bone resorption (mean −0.43 mm), with significantly greater effects in adults than adolescents (low-certainty evidence). The clinical significance may vary with baseline alveolar thickness. Pre-treatment tomographic assessment is recommended for adult patients when clinically justified.
3. **En-masse retraction** results in faster space closure than two-step retraction (mean reduction 4.2 months) with comparable anchorage loss, incisor retraction, and root resorption risk (moderate-certainty evidence).
4. **Root resorption** incidence is 12.4% (13.2% for >2 mm threshold, 11.4% for ≥¼ root length threshold), with higher risk in maxillary incisors (OR 2.34) (moderate-certainty evidence).
5. **Extraction versus non-extraction** protocols demonstrate comparable long-term stability for overjet, overbite, and incisor irregularity (low-certainty evidence), suggesting that extraction decisions should be based on treatment objectives rather than stability concerns.
6. **Patient age** significantly influences bone remodeling capacity, with adults showing greater palatal bone resorption than adolescents following incisor retraction.
7. **Clinical decision-making** should consider patient-specific factors including age, alveolar bone morphology, malocclusion pattern, and appliance selection. The evidence-based framework and clinical algorithm provided by this meta-analysis support individualized treatment planning.

## Supporting information

Supplementary File 1: R Code for Meta-Analyses

Supplementary File 2: Data Extraction Form Template

Supplementary Table 1: Full Search Strategy for Each Database

Supplementary Table 2: PRISMA 2020 Checklist

Supplementary Table 3: Full List of Excluded Studies with Reasons

## SUPPLEMENTARY MATERIALS

All extracted data, analytic code (R scripts), and data collection templates are provided as supplementary materials accompanying this article. All materials are publicly available within the supplementary files of this publication.

**Supplementary Table 1.** Full search strategy for each database

**Supplementary Table 2.** PRISMA 2020 checklist

**Supplementary Table 3.** Full list of excluded studies with reasons (extended version)

**Supplementary File 1.** R code for meta-analyses with reproducibility notes

**Supplementary File 2.** Data extraction form template

## Author Contributions

**Maen Mahfouz:** Conceptualization, methodology, investigation, writing – original draft preparation, writing – review and editing, supervision. **Eman Alzaben:** Investigation, writing – original draft preparation, writing – review and editing.

## Data Availability

All data extracted from included studies, analytic code (R scripts), and data collection templates are provided as supplementary materials accompanying this article. The data that support the findings of this study are available from the corresponding author upon reasonable request. Original source data are available in the published articles cited in the references.

