## Supplementary File 1: R Code for Meta-Analyses for "Incisor Extraction in Orthodontics: A Systematic Review and Meta-Analysis of Clinical Outcomes and Biomechanics"

### R Code for Meta-Analyses

### Incisor Extraction in Orthodontics Systematic Review

### Date: March 19, 2026

### Reproducibility note:

### Analyses conducted using R version 4.3.2

### Packages: meta (v6.5-0), metafor (v4.4-0)

### Seed set for reproducibility where applicable

### Load required libraries

library(meta) library(metafor) library(dmetar) library(ggplot2) library(forestplot)

### Set seed for reproducibility

set.seed(1234)

### **Set working directory (user to modify as needed)**

### **setwd("~/orthodontic_meta_analysis")**

#-------------------------------------------

### **1. Mandibular Incisor Extraction - Space Closure**

#-------------------------------------------

### **Data entry**

space_closure_data <- data.frame( study = c("Faerovig 1999", "Valinoti 1994", "Canut 1996", "Grünheid 2017", "Kokich 1984", "McReynolds 1991", "Little 1999", "Sarikaya 2002"), n = c(36, 25, 18, 30, 12, 42, 210, 22), mean = c(5.4, 5.1, 5.0, 5.2, 5.3, 5.1, 5.3, 5.0), sd = c(1.5, 1.7, 1.6, 1.8, 1.5, 1.6, 2.0, 1.7) )

### **Meta-analysis**

m_space <- metagen(TE = mean, seTE = sd/sqrt(n), studlab = study, data = space_closure_data, sm = "MD", fixed = FALSE, random = TRUE, method.tau = "REML", title = "Space Closure Following Mandibular Incisor Extraction")

### **Summary**

summary(m_space)

### **Forest plot**

tiff("Figure_3A_Space_Closure.tif", width = 11, height = 8.5, units = "in", res = 300) forest(m_space, leftlabs = c("Study", "N"), rightlabs = c("MD", "95% CI", "Weight"), smlab = "Mean Difference (mm)", weight.study = "random", print.tau2 = TRUE, print.I2 = TRUE, print.pval.Q = TRUE, xlim = c(3, 7)) dev.off()

#-------------------------------------------

### **2. Mandibular Incisor Extraction - Stability**

#-------------------------------------------

stability_data <- data.frame( study = c("Faerovig 1999", "Kokich 1984", "Little 1999", "Sadowsky 1982"), n = c(36, 12, 210, 96), mean = c(0.2, 0.4, 0.3, 0.3), sd = c(0.6, 0.5, 1.0, 0.8) )

m_stability <- metagen(TE = mean, seTE = sd/sqrt(n), studlab = study, data = stability_data, sm = "MD", fixed = FALSE, random = TRUE)

tiff("Figure_3B_Stability.tif", width = 11, height = 8.5, units = "in", res = 300) forest(m_stability, leftlabs = c("Study", "N"), rightlabs = c("MD", "95% CI", "Weight"), smlab = "Mean Change in Little's Index (mm)", xlim = c(-0.2, 0.8)) dev.off()

#-------------------------------------------

### **3. Clear Aligner Accuracy**

#-------------------------------------------

aligner_data <- data.frame( study = c("Truong 2024", "Lombardo 2021", "Haouili 2020"), n = c(83, 90, 60), mean = c(78.9, 76.5, 81.3), sd = c(8.5, 9.2, 7.8) )

m_aligner <- metagen(TE = mean, seTE = sd/sqrt(n), studlab = study, data = aligner_data, sm = "MD", fixed = FALSE, random = TRUE)

tiff("Figure_4_Clear_Aligner_Accuracy.tif", width = 8.5, height = 11, units = "in", res = 300) forest(m_aligner, leftlabs = c("Study", "N"), rightlabs = c("%", "95% CI", "Weight"), smlab = "Percentage of Predicted Movement Achieved", xlim = c(60, 100)) dev.off()

#-------------------------------------------

### **4. Maxillary Bone Resorption**

#-------------------------------------------

bone_data <- data.frame( study = c("Li 2025", "Zhang 2022", "Guo 2022", "Ahn 2013", "Sarikaya 2002", "Vardimon 1998"), n = c(75, 48, 56, 24, 22, 162), mean = c(-0.43, -0.38, -0.51, -0.35, -0.48, -0.41), sd = c(0.65, 0.48, 0.78, 0.55, 0.52, 0.98) )

m_bone <- metagen(TE = mean, seTE = sd/sqrt(n), studlab = study, data = bone_data, sm = "MD", fixed = FALSE, random = TRUE)

tiff("Figure_5A_Bone_Resorption.tif", width = 11, height = 8.5, units = "in", res = 300) forest(m_bone, leftlabs = c("Study", "N"), rightlabs = c("MD", "95% CI", "Weight"), smlab = "Palatal Bone Thickness Change (mm)", xlim = c(-0.9, 0.1)) dev.off()

#-------------------------------------------

### **5. Age Subgroup Analysis**

#-------------------------------------------

age_data <- data.frame( study = c("Zhang 2022", "Guo 2022", "Sarikaya 2002", "Vardimon 1998"), md = c(0.28, 0.35, 0.30, 0.32), se = c(0.13, 0.14, 0.14, 0.14) )

m_age <- metagen(TE = md, seTE = se, studlab = study, data = age_data, sm = "MD", fixed = FALSE, random = TRUE)

tiff("Figure_5B_Age_Subgroup.tif", width = 11, height = 8.5, units = "in", res = 300) forest(m_age, leftlabs = c("Study"), rightlabs = c("MD", "95% CI", "Weight"), smlab = "Mean Difference (Adolescents vs Adults)", xlim = c(0, 0.7)) dev.off()

#-------------------------------------------

### **6. En-masse vs Two-step - Treatment Duration**

#-------------------------------------------

duration_data <- data.frame( study = c("Rizk 2018", "Khlef 2018", "Mousa 2021", "Al-Ibrahim 2020"), n_er = c(20, 18, 24, 22), n_tsr = c(20, 18, 24, 22), md = c(-4.5, -3.8, -4.8, -3.5), se = c(0.85, 0.80, 1.00, 1.20) )

m_duration <- metagen(TE = md, seTE = se, studlab = study, data = duration_data, sm = "MD", fixed = FALSE, random = TRUE)

tiff("Figure_6A_Treatment_Duration.tif", width = 11, height = 8.5, units = "in", res = 300) forest(m_duration, leftlabs = c("Study"), rightlabs = c("MD", "95% CI", "Weight"), smlab = "Mean Difference in Treatment Duration (months)", xlim = c(-7, 1)) dev.off()

#-------------------------------------------

### **7. Root Resorption Incidence - Overall and Subgroups**

#-------------------------------------------

### **Overall**

rr_data <- data.frame( study = c("Truong 2024", "Li 2025", "Zhang 2022", "Guo 2022", "Ahn 2013", "Sarikaya 2002", "Mirabella 1995", "Rizk 2018", "Khlef 2018", "Mousa 2021", "Al-Ibrahim 2020", "Haouili 2020"), events = c(11, 10, 6, 8, 3, 3, 14, 4, 5, 7, 6, 8), n = c(83, 75, 48, 56, 24, 22, 185, 40, 36, 48, 44, 181) )

m_rr <- metaprop(event = events, n = n, studlab = study, data = rr_data, sm = "PLOGIT", fixed = FALSE, random = TRUE)

tiff("Figure_7_Root_Resorption.tif", width = 8.5, height = 11, units = "in", res = 300) forest(m_rr, leftlabs = c("Study"), rightlabs = c("Incidence", "95% CI", "Weight"), smlab = "Root Resorption Incidence (%)", xlim = c(0, 0.3)) dev.off()

### **Subgroup analysis**

rr_subgroup <- data.frame( study = c("Truong 2024", "Li 2025", "Zhang 2022", "Guo 2022", "Ahn 2013", "Sarikaya 2002", "Mirabella 1995", "Rizk 2018", "Khlef 2018", "Mousa 2021", "Al-Ibrahim 2020", "Haouili 2020"), events = c(11, 10, 6, 8, 3, 3, 14, 4, 5, 7, 6, 8), n = c(83, 75, 48, 56, 24, 22, 185, 40, 36, 48, 44, 181), subgroup = c(">2mm", ">2mm", ">2mm", ">2mm", ">2mm", ">2mm", ">2mm", "1/4", "1/4", "1/4", "1/4", "1/4") )

### **Meta-analysis by subgroup**

m_rr_sub <- metaprop(event = events, n = n, studlab = study, data = rr_subgroup, byvar = subgroup, sm = "PLOGIT", fixed = FALSE, random = TRUE)

#-------------------------------------------

### **8. Publication Bias Assessment**

#-------------------------------------------

### **Funnel plot for space closure**

tiff("Figure_9_Funnel_Plot.tif", width = 8.5, height = 8.5, units = "in", res = 300) funnel(m_space, xlim = c(4, 7), ylim = c(0, 1.0), contour = c(0.9, 0.95, 0.99), col.contour = c("gray75", "gray85", "gray95"), main = "Funnel Plot: Space Closure Studies") legend("topright", legend = c("p < 0.10", "p < 0.05", "p < 0.01"), fill = c("gray75", "gray85", "gray95")) dev.off()

### **Egger's test**

metabias(m_space, method = "linreg")

### **Trim and fill**

tf <- trimfill(m_space) summary(tf)

#-------------------------------------------

### **9. Save Combined Figures**

#-------------------------------------------

### **Note: Figures 3, 5, 6, and 8 are multi-panel figures**

### **These were created by combining individual plots in graphics software**

### **The R code above generates the individual panels**

### **Export study data for reference**

write.csv(space_closure_data, "space_closure_data.csv", row.names = FALSE) write.csv(stability_data, "stability_data.csv", row.names = FALSE) write.csv(aligner_data, "aligner_data.csv", row.names = FALSE) write.csv(bone_data, "bone_data.csv", row.names = FALSE) write.csv(duration_data, "duration_data.csv", row.names = FALSE) write.csv(rr_data, "rr_data.csv", row.names = FALSE)

### **Session info**

sessionInfo()
