## Supplementary File 2: Data Extraction Form Template for "Incisor Extraction in Orthodontics: A Systematic Review and Meta-Analysis of Clinical Outcomes and Biomechanics"

### **Supplementary File 2: Data Extraction Form Template SYSTEMATIC REVIEW DATA EXTRACTION FORM**

##### **Incisor Extraction in Orthodontics: A Systematic Review and Meta-Analysis**

| **STUDY ID** |
| --- |
| **Reviewer** |
| **Date** |

##### **STUDY CHARACTERISTICS**

| **Field** | **Entry** |
| --- | --- |
| First Author |  |
| Year |  |
| Country |  |
| Study Design | □ RCT □ Prospective cohort □ Retrospective cohort |
| Setting | □ University □ Hospital □ Private practice □ Mixed |
| Sample Size (total) |  |
| Sample Size (extraction group) |  |
| Sample Size (control/non-extraction) |  |
| Follow-up duration | months / years |
| Funding source |  |
| Conflicts of interest reported | □ Yes □ No □ Not stated |

##### **PARTICIPANT CHARACTERISTICS**

| **Field** | **Entry** |
| --- | --- |
| Age (mean ± SD) | Mean ______ (SD ______) |
| Age (range) | ______ - ______ |
| Sex (male) |  |
| Sex (female) |  |
| Sex (not specified) |  |
| Malocclusion type |  |
| Extraction pattern |  |
| Inclusion criteria |  |
| Exclusion criteria |  |

##### **INTERVENTION DETAILS**

| **Field** | **Entry** |
| --- | --- |
| **Extraction site** |  |
| □ Mandibular incisor |  |
| □ Maxillary incisor |  |
| □ Premolar (maxillary) |  |
| □ Premolar (mandibular) |  |
| □ Mixed |  |
| **Appliance type** |  |
| □ Fixed (type: _______________) |  |
| □ Clear aligners (system: _______________) |  |
| **Retraction mechanics** |  |
| □ En-masse |  |
| □ Two-step |  |
| □ Not applicable |  |
| **Anchorage** |  |
| □ Conventional |  |
| □ TAD-assisted |  |
| □ Other: _______________ |  |
| Force magnitude | _______________ g |
| Treatment duration | _______________ months |
| Number of patients completing treatment |  |

##### **OUTCOME MEASURES**

| **Field** | **Entry** |
| --- | --- |
| **Assessment method** |  |
| □ CBCT |  |
| □ CT |  |
| □ Cephalometric |  |
| □ Model analysis |  |
| **Blinded assessment** | □ Yes □ No □ Not specified |
| **Calibration performed** | □ Yes □ No □ Not specified |
| **Intra-examiner reliability reported** | □ Yes □ No |
| **Inter-examiner reliability reported** | □ Yes □ No |

##### **PRIMARY OUTCOMES REPORTED**

###### **□ Space closure amount**

| Mean | ______ |
| --- | --- |
| SD | ______ |
| n | ______ |
| Measurement method |  |

###### **□ Little's Irregularity Index**

| Pre-treatment mean (SD) | ______ (______) |
| --- | --- |
| Post-treatment mean (SD) | ______ (______) |
| Change mean (SD) | ______ (______) |
| n | ______ |

###### **□ Root resorption incidence**

| Events | ______ |
| --- | --- |
| n | ______ |
| Definition used | □ >2 mm □ ≥¼ root length □ Other: _______________ |
| Assessment method | □ Periapical □ Panoramic □ CBCT |

###### **□ Bone thickness change (palatal)**

|  | Central Incisors | Lateral Incisors |
| --- | --- | --- |
| Mean (SD) | ______ (______) | ______ (______) |
| n | ______ | ______ |
| Measurement level | □ Cervical □ Mid-root □ Apical | □ Cervical □ Mid-root □ Apical |

###### **□ Bone thickness change (labial)**

|  | Central Incisors | Lateral Incisors |
| --- | --- | --- |
| Mean (SD) | ______ (______) | ______ (______) |
| n | ______ | ______ |

###### **□ Treatment duration**

|  | ER Group | TSR Group |
| --- | --- | --- |
| Mean (SD) | ______ (______) | ______ (______) |
| n | ______ | ______ |

###### **□ Anchorage loss**

| Mean | ______ |
| --- | --- |
| SD | ______ |
| n | ______ |
| Measurement method |  |

###### **□ Incisor retraction amount**

| Mean | ______ |
| --- | --- |
| SD | ______ |
| n | ______ |

###### **□ Overjet relapse**

|  | Extraction Group | Non-extraction Group |
| --- | --- | --- |
| Mean (SD) | ______ (______) | ______ (______) |
| n | ______ | ______ |

###### **□ Overbite relapse**

|  | Extraction Group | Non-extraction Group |
| --- | --- | --- |
| Mean (SD) | ______ (______) | ______ (______) |
| n | ______ | ______ |

###### **□ Incisor irregularity relapse**

|  | Extraction Group | Non-extraction Group |
| --- | --- | --- |
| Mean (SD) | ______ (______) | ______ (______) |
| n | ______ | ______ |

##### **RESULTS FOR META-ANALYSIS (if applicable)**

###### **Continuous outcomes (extraction group)**

| Outcome | Mean | SD | N |
| --- | --- | --- | --- |
| 1. |  |  |  |
| 2. |  |  |  |
| 3. |  |  |  |

###### **Continuous outcomes (control group)**

| Outcome | Mean | SD | N |
| --- | --- | --- | --- |
| 1. |  |  |  |
| 2. |  |  |  |
| 3. |  |  |  |

###### **Dichotomous outcomes**

| Outcome | Events | Total N |
| --- | --- | --- |
| 1. |  |  |
| 2. |  |  |
| 3. |  |  |

##### **RISK OF BIAS ASSESSMENT**

###### **For RCTs (RoB 2.0)**

| Domain | Low Risk | Some Concerns | High Risk |
| --- | --- | --- | --- |
| Randomization process | □ | □ | □ |
| Deviations from intended interventions | □ | □ | □ |
| Missing outcome data | □ | □ | □ |
| Measurement of outcome | □ | □ | □ |
| Selection of reported result | □ | □ | □ |
| **Overall bias** | □ | □ | □ |

###### **For observational studies (ROBINS-I)**

| Domain | Low | Moderate | Serious | Critical |
| --- | --- | --- | --- | --- |
| Bias due to confounding | □ | □ | □ | □ |
| Bias in selection of participants | □ | □ | □ | □ |
| Bias in classification of interventions | □ | □ | □ | □ |
| Bias due to deviations from interventions | □ | □ | □ | □ |
| Bias due to missing data | □ | □ | □ | □ |
| Bias in measurement of outcomes | □ | □ | □ | □ |
| Bias in selection of reported result | □ | □ | □ | □ |
| **Overall bias** | □ | □ | □ | □ |

##### **ADDITIONAL NOTES**

**Form completed by:** ___________________________ **Date:** ___________________________
