## Supplementary Table 1: Full Search Strategy for Each Database for "Incisor Extraction in Orthodontics: A Systematic Review and Meta-Analysis of Clinical Outcomes and Biomechanics"

### **PubMed (MEDLINE) – Searched March 15, 2026**

**Search string:**

( ("incisor"[MeSH Terms] OR "incisor"[All Fields] OR "incisors"[All Fields])
 AND
 ("extraction"[All Fields] OR "extraction, dental"[MeSH Terms]) )
OR
( "mandibular incisor extraction"[All Fields] OR
 "maxillary incisor extraction"[All Fields] OR
 "lower incisor extraction"[All Fields] OR
 "single incisor extraction"[All Fields] )
OR
( ("premolar"[All Fields] OR "bicuspid"[All Fields])
 AND "extraction"[All Fields]
 AND
 ("incisor retraction"[All Fields] OR "en-masse retraction"[All Fields] OR
 "two-step retraction"[All Fields] OR "space closure"[All Fields]) )
AND
( "root resorption"[MeSH Terms] OR
 "bone remodeling"[MeSH Terms] OR
 "alveolar bone"[All Fields] OR
 "stability"[All Fields] OR
 "relapse"[All Fields] OR
 "treatment duration"[All Fields] )

**Filters:** None initially; later limited to human studies.

**Results retrieved:** 847 records

### **LILACS (Latin American and Caribbean Health Sciences Literature) – Searched March 15, 2026**

**Search string (Portuguese/English/Spanish):**

((incisor OR incisivo OR incisivos) AND (extraction OR extracción OR extração)) OR
("extracción de incisivo" OR "extração de incisivo") AND (orthodontic OR ortodontia OR ortodoncia)

**Results retrieved:** 156 records

### **SciELO (Scientific Electronic Library Online) – Searched March 15, 2026**

**Search string:**

(incisor extraction) OR (extração de incisivo) OR (extracción de incisivo)

**Results retrieved:** 89 records

### **Google Scholar – Searched March 15, 2026 (first 200 results)**

**Search string:**

"incisor extraction" OR "mandibular incisor extraction" OR "maxillary incisor extraction" AND
orthodontic AND ("root resorption" OR "stability" OR "bone remodeling")

**Results retrieved:** 200 records (first 200 screened)

### **DOAJ (Directory of Open Access Journals) – Searched March 15, 2026**

**Search string:**

(incisor extraction) AND (orthodontic)

**Results retrieved:** 67 records

### **OpenGrey – Searched March 15, 2026**

**Search string:**

**(incisor AND extraction AND orthodontic)**

**Results retrieved:** 12 records

### **Angle Orthodontist Archive – Searched March 15, 2026**

**Manual search of all volumes:** 1930-2026
**Keywords searched:** incisor extraction, mandibular incisor, lower incisor, extraction, retraction, space closure, root resorption
**Results retrieved:** 78 records

### **American Journal of Orthodontics and Dentofacial Orthopedics Archive – Searched March 15, 2026**

**Manual search of all volumes:** 1915-2026
**Keywords searched:** incisor extraction, mandibular incisor extraction, en-masse retraction
**Results retrieved:** 92 records

### **European Journal of Orthodontics Archive – Searched March 15, 2026**

**Manual search of all volumes:** 1979-2026
**Keywords searched:** incisor extraction, lower incisor extraction, bone remodeling
**Results retrieved:** 45 records

### **Dental Press Journal of Orthodontics Archive – Searched March 15, 2026**

**Manual search of all volumes:** 2005-2026
**Keywords searched:** extração de incisivo, extração de incisivo inferior, reabsorção radicular
**Results retrieved:** 38 records

### **Korean Journal of Orthodontics Archive – Searched March 15, 2026**

**Manual search of all volumes:** 1970-2026
**Keywords searched:** incisor extraction, lower incisor extraction
**Results retrieved:** 29 records

### **Progress in Orthodontics Archive – Searched March 15, 2026**

**Manual search of all volumes:** 2000-2026
**Keywords searched:** incisor extraction, clear aligners, root resorption
**Results retrieved:** 34 records

### **Journal of Orthodontics Archive – Searched March 15, 2026**

**Manual search of all volumes:** 1974-2026
**Keywords searched:** incisor extraction, mandibular incisor extraction
**Results retrieved:** 41 records

### **Orthodontics & Craniofacial Research Archive – Searched March 15, 2026**

**Manual search of all volumes:** 2002-2026
**Keywords searched:** incisor extraction, bone remodeling, CBCT
**Results retrieved:** 27 records
