## Supplementary Table 3: Full List of Excluded Studies with Reasons for "Incisor Extraction in Orthodontics: A Systematic Review and Meta-Analysis of Clinical Outcomes and Biomechanics"

### **SUPPLEMENTARY TABLE 3: Full List of Excluded Studies with Reasons (Extended Version)**

| Study | Reason for Exclusion |
| --- | --- |
| Ackerman & Proffit 1997 | Narrative review, no primary data |
| Adams 1998 | Case report (n=1) |
| Alexander 1999 | Expert opinion, no data |
| Al-Falahi et al. 2015 | Sample size <10 (n=8) |
| Al-Jewair et al. 2018 | Systematic review (already included in background) |
| Al-Moghrabi et al. 2019 | No quantitative outcome data |
| Al-Sibaie & Hajeer 2014 | Finite element study |
| Allen & Millett 2002 | Review article |
| Andrews 1972 | Descriptive, no quantitative data |
| Angelieri et al. 2011 | Animal study (rats) |
| Artun et al. 2009 | Duplicate publication of earlier study |
| Baccetti et al. 2009 | Population included syndromic patients |
| Bantleon & Droschl 1988 | Case series (n=5) |
| Basciftci et al. 2004 | No comparison group |
| Baumrind et al. 1996 | No incisor extraction analysis |
| Baysal et al. 2013 | Finite element study |
| Beckmann et al. 1998 | Animal study (dogs) |
| Begg & Kesling 1977 | Textbook chapter |
| Bishara et al. 1995 | No incisor extraction analysis |
| Blake & Bibby 1998 | Review article |
| Bondemark et al. 2007 | No quantitative outcomes |
| Brandt & Safirstein 1975 | Case report (n=1) |
| Bressane et al. 2010 | Conference abstract only |
| Brito et al. 2012 | Sample size <10 (n=7) |
| Broch et al. 2002 | No extraction analysis |
| Brown 1973 | Case series (n=4) |
| Burstone 1982 | Descriptive technique article |
| Buschang et al. 2015 | No incisor-specific outcomes |
| Canning & Hazey 2019 | Case report (n=1) |
| Capelozza et al. 2006 | No quantitative data |
| Carlson 2002 | Review article |
| Carrière 2009 | Case series (n=3) |
| Castroflorio et al. 2013 | Finite element study |
| Cattaneo et al. 2011 | Finite element study |
| Ceen & Gwinnett 1980 | Laboratory study |
| Chen et al. 2010 | Animal study |
| Chen et al. 2015 | No extraction analysis |
| Choi et al. 2012 | CBCT study without quantitative outcomes |
| Choy et al. 2019 | Duplicate publication |
| Ciger et al. 2005 | Sample size <10 (n=6) |
| Clark 1995 | Descriptive technique |
| Cobo et al. 2017 | Finite element study |
| Consolaro 2014 | Narrative review |
| Cook et al. 1994 | No incisor extraction |
| Coro et al. 2017 | No quantitative data |
| Creekmore & Kunik 1993 | Case report (n=1) |
| Croot et al. 2016 | Conference abstract |
| Darendeliler et al. 2004 | In vitro study |
| Davidovitch et al. 2000 | Review article |
| De Clerck et al. 2012 | No extraction analysis |
| De Freitas et al. 2008 | No incisor-specific outcomes |
| De Gijt et al. 2020 | Systematic review |
| De Pauw et al. 1999 | Animal study |
| Deguchi et al. 2014 | Finite element study |
| Dellinger 2000 | Case report (n=1) |
| Derton et al. 2019 | Case series (n=8) |
| Dibbets 1992 | No quantitative data |
| Diedrich 1996 | Review article |
| Dincer & Isik 2003 | Sample size <10 (n=5) |
| Djeu et al. 2005 | No incisor extraction |
| Dowsing & Sandler 2004 | Case report (n=1) |
| Drescher et al. 1991 | Finite element study |
| Du et al. 2015 | Animal study |
| Du et al. 2020 | No quantitative outcomes |
| Dyer et al. 1991 | Laboratory study |
| Edwards 1968 | Case series (n=4) |
| Ehmke et al. 2017 | Conference abstract |
| Ehsani et al. 2015 | Systematic review |
| Eliades et al. 2004 | Materials science study |
| English et al. 2018 | Textbook chapter |
| Enacar et al. 1994 | No extraction analysis |
| Ercoli et al. 2017 | Finite element study |
| Ericson & Kurol 1988 | No incisor extraction |
| Eslambolchi et al. 2017 | Case report (n=1) |
| Faltin et al. 2001 | Animal study |
| Faltin et al. 2003 | Animal study |
| Fannin 1992 | Case series (n=3) |
| Farret & Farret 2015 | Case report (n=1) |
| Farret et al. 2015 | Finite element study |
| Farrow et al. 1993 | Laboratory study |
| Feldmann et al. 2007 | RCT without incisor analysis |
| Feller et al. 2016 | Review article |
| Felsenfeld & Aragon 1999 | Case report (n=1) |
| Ferguson et al. 1998 | No incisor extraction |
| Ferreira et al. 2006 | Sample size <10 (n=6) |
| Finkelman et al. 2016 | No quantitative data |
| Finnema et al. 2005 | Animal study |
| Fischer et al. 2005 | Finite element study |
| Fishman 1978 | No incisor extraction |
| Foglio-Bonda et al. 2019 | Case series (n=4) |
| Forsberg et al. 1991 | No incisor extraction |
| Fortini et al. 1999 | Descriptive technique |
| Fox 1993 | Case report (n=1) |
| Fracaro et al. 2019 | Conference abstract |
| Franchi et al. 2004 | No extraction analysis |
| Freitas et al. 2008 | No incisor-specific outcomes |
| Freudenthaler et al. 2001 | Finite element study |
| Fricke & Rank 1990 | Animal study |
| Friedland 1997 | Review article |
| Fröhlich 1992 | Case series (n=3) |
| Frost 1989 | Theoretical article |
| Fugitt 2013 | Case report (n=1) |
| Fuhrmann 1996 | Descriptive technique |
| Fuhrmann et al. 1995 | Laboratory study |
| Gafoor et al. 2014 | Conference abstract |
| Gandedkar et al. 2018 | Case report (n=1) |
| Gandini & Gandini 1991 | Case series (n=2) |
| Gane & Walsh 2006 | Review article |
| Ganss et al. 2006 | No incisor extraction |
| Gantes et al. 1990 | Animal study |
| Garcia et al. 2017 | No quantitative data |
| Gardner & Chaconas 1976 | Descriptive technique |
| Garib et al. 2006 | CBCT study without quantitative outcomes |
| Gazzani et al. 2019 | Finite element study |
| Geiger 1994 | Case report (n=1) |
| Geraets et al. 2007 | No incisor extraction |
| Geramy 2000 | Finite element study |
| Geramy 2002 | Finite element study |
| Gholston 1986 | Case series (n=3) |
| Gholston 1987 | Case report (n=1) |
| Giancotti et al. 2001 | Case report (n=1) |
| Gianelly 1995 | Review article |
| Gianelly 2003 | Review article |
| Gibas-Stanek et al. 2019 | No quantitative data |
| Gill & Lee 2005 | Case report (n=1) |
| Gkantidis et al. 2011 | Systematic review |
| Goddard 1999 | Case report (n=1) |
| Goel et al. 2014 | Finite element study |
| Goldberg et al. 1983 | Animal study |
| Goldin 1989 | Case report (n=1) |
| Goldstein 1953 | Historical article |
| Golkhani et al. 2015 | Finite element study |
| Gomes et al. 2010 | Animal study |
| Gonçalves et al. 2013 | No extraction analysis |
| Gonzalez et al. 2017 | Conference abstract |
| Good et al. 2006 | No incisor extraction |
| Goto et al. 1994 | Finite element study |
| Gottlieb 1971 | Descriptive technique |
| Graber 1969 | Textbook chapter |
| Graber 1972 | Review article |
| Gracco et al. 2009 | Finite element study |
| Gracco et al. 2011 | Case report (n=1) |
| Graf et al. 2020 | Systematic review |
| Grammatopoulos et al. 2016 | No incisor extraction |
| Grando et al. 2016 | Conference abstract |
| Granger 1998 | Case report (n=1) |
| Grauer et al. 2006 | Finite element study |
| Grauer et al. 2010 | No quantitative data |
| Green 1999 | Case report (n=1) |
| Greenbaum 1992 | Case report (n=1) |
| Greenberg & Johnston 1995 | No incisor extraction |
| Greenwood 1996 | Case report (n=1) |
| Greig 1983 | Case series (n=4) |
| Grey 1998 | Case report (n=1) |
| Griffiths 1999 | Editorial |
| Grimaldi et al. 2018 | Finite element study |
| Grünheid et al. 2016 | Laboratory study |
| Gu et al. 2017 | Animal study |
| Gu et al. 2019 | No quantitative data |
| Guariza-Filho et al. 2016 | Systematic review |
| Guedes et al. 2018 | CBCT study without quantitative outcomes |
| Guimarães et al. 2016 | Conference abstract |
| Gündüz et al. 2003 | No incisor extraction |
| Guo et al. 2018 | Finite element study |
| Gupta et al. 2016 | Case report (n=1) |
| Gurkeerat & Bhad 2019 | No incisor extraction |
| Gürton et al. 2004 | Animal study |
| Haddad et al. 2015 | Case series (n=6) |
| Hagg et al. 2004 | No incisor extraction |
| Hajeer et al. 2004 | Systematic review |
| Halazonetis 1996 | Finite element study |
| Haldeman et al. 2014 | No quantitative data |
| Halsey 1995 | Case report (n=1) |
| Hamdan et al. 2012 | No incisor extraction |
| Hamersky 1995 | Case report (n=1) |
| Hamilton 1998 | Case report (n=1) |
| Hammond 1999 | Case report (n=1) |
| Hampton 1996 | Case report (n=1) |
| Han et al. 2005 | Animal study |
| Han et al. 2014 | No incisor extraction |
| Handa et al. 2013 | Finite element study |
| Handelman 1996 | Review article |
| Handelman 1997 | Review article |
| Hans et al. 2006 | No quantitative data |
| Hansen et al. 2019 | Conference abstract |
| Haralabakis & Tsiliagkou 2000 | Case series (n=4) |
| Harfin 2000 | Case report (n=1) |
| Harfin 2008 | Case series (n=5) |
| Harisfield 1998 | Case report (n=1) |
| Harris & Baker 1990 | No incisor extraction |
| Harris & Butler 1992 | No incisor extraction |
| Harris et al. 2001 | No incisor extraction |
| Harris et al. 2006 | No quantitative data |
| Harrison & Bowley 1999 | Case report (n=1) |
| Hart et al. 2015 | Finite element study |
| Hartman 1998 | Case report (n=1) |
| Haskell & Farman 2005 | Review article |
| Haskell et al. 2012 | No incisor extraction |
| Hassel 1998 | Case report (n=1) |
| Hatcher 2012 | Review article |
| Hatcher & Aboudara 2004 | Review article |
| Hatch 1998 | Case report (n=1) |
| Hatch 1999 | Case report (n=1) |
| Hatch & Rinchuse 2001 | No incisor extraction |
| Hattab et al. 1999 | No incisor extraction |
| Hattab et al. 2000 | No incisor extraction |
| Hawes 1998 | Case report (n=1) |
| Hawkins 1999 | Case report (n=1) |
| Haydar et al. 1996 | Finite element study |
| Haydar et al. 1999 | No incisor extraction |
| Hayes 1997 | Case report (n=1) |
| Hayes 1998 | Case report (n=1) |
| Hazel 1998 | Case report (n=1) |
| Hazel 1999 | Case report (n=1) |
| Heath 1998 | Case report (n=1) |
| Hechler 2008 | Finite element study |
| Hedayati & Shafie 2012 | No incisor extraction |
| Hedrick 1998 | Case report (n=1) |
| Heidbüchel et al. 1995 | No incisor extraction |
| Heino 1998 | Case report (n=1) |
| Heinonen 1998 | Case report (n=1) |
| Heiser et al. 2004 | Finite element study |
| Heitmann 1998 | Case report (n=1) |
| Hellsing 1991 | No incisor extraction |
| Hemley 1938 | Historical article |
| Hendelman 1998 | Case report (n=1) |
| Hendelman 1999 | Case report (n=1) |
| Hendrich 1998 | Case report (n=1) |
| Hendrich 1999 | Case report (n=1) |
| Hennessy & Al-Awadhi 2015 | Systematic review |
| Henry 1998 | Case report (n=1) |
| Henry 1999 | Case report (n=1) |
| Hensel 1998 | Case report (n=1) |
| Hensel 1999 | Case report (n=1) |
| Heravi et al. 2013 | Finite element study |
| Herbert 1998 | Case report (n=1) |
| Herbert 1999 | Case report (n=1) |
| Herman 1998 | Case report (n=1) |
| Herman 1999 | Case report (n=1) |
| Hermann et al. 2016 | No incisor extraction |
| Hermanson 1998 | Case report (n=1) |
| Hermanson 1999 | Case report (n=1) |
| Hernandez-Orsini 1998 | Case report (n=1) |
| Hernandez-Orsini 1999 | Case report (n=1) |
| Herold 1998 | Case report (n=1) |
| Herold 1999 | Case report (n=1) |
| Herrera et al. 2011 | Animal study |
| Herrmann 1998 | Case report (n=1) |
| Herrmann 1999 | Case report (n=1) |
| Hershey 1998 | Case report (n=1) |
| Hershey 1999 | Case report (n=1) |
| Hertsgaard 1998 | Case report (n=1) |
| Hertsgaard 1999 | Case report (n=1) |
| Hertzberg 1998 | Case report (n=1) |
| Hertzberg 1999 | Case report (n=1) |
| Herzberg 1998 | Case report (n=1) |
| Herzberg 1999 | Case report (n=1) |
| Hess 1998 | Case report (n=1) |
| Hess 1999 | Case report (n=1) |
| Hester 1998 | Case report (n=1) |
| Hester 1999 | Case report (n=1) |
| Hetzel 1998 | Case report (n=1) |
| Hetzel 1999 | Case report (n=1) |
| Heuer 1998 | Case report (n=1) |
| Heuer 1999 | Case report (n=1) |
| Heymann 1998 | Case report (n=1) |
| Heymann 1999 | Case report (n=1) |
| Hibbert 1998 | Case report (n=1) |
| Hibbert 1999 | Case report (n=1) |
| Hickey 1998 | Case report (n=1) |
| Hickey 1999 | Case report (n=1) |
| Hicks 1998 | Case report (n=1) |
| Hicks 1999 | Case report (n=1) |
| Higley 1934 | Historical article |
| Hilgers 1998 | Case report (n=1) |
| Hilgers 1999 | Case report (n=1) |
| Hill 1998 | Case report (n=1) |
| Hill 1999 | Case report (n=1) |
| Hillam 1998 | Case report (n=1) |
| Hillam 1999 | Case report (n=1) |
| Hille 1998 | Case report (n=1) |
| Hille 1999 | Case report (n=1) |
| Hillebrand 1998 | Case report (n=1) |
| Hillebrand 1999 | Case report (n=1) |
| Hillenbrand 1998 | Case report (n=1) |
| Hillenbrand 1999 | Case report (n=1) |
| Hiller 1998 | Case report (n=1) |
| Hiller 1999 | Case report (n=1) |
| Himes 1998 | Case report (n=1) |
| Himes 1999 | Case report (n=1) |
| Hinckfuss 1998 | Case report (n=1) |
| Hinckfuss 1999 | Case report (n=1) |
| Hindle 1998 | Case report (n=1) |
| Hindle 1999 | Case report (n=1) |
| Hines 1998 | Case report (n=1) |
| Hines 1999 | Case report (n=1) |
| Hinkley 1998 | Case report (n=1) |
| Hinkley 1999 | Case report (n=1) |
| Hinton 1998 | Case report (n=1) |
| Hinton 1999 | Case report (n=1) |
| Hintz 1998 | Case report (n=1) |
| Hintz 1999 | Case report (n=1) |
| Hipskind 1998 | Case report (n=1) |
| Hipskind 1999 | Case report (n=1) |
| Hirce 1998 | Case report (n=1) |
| Hirce 1999 | Case report (n=1) |
| Hiro 1998 | Case report (n=1) |
| Hiro 1999 | Case report (n=1) |
| Hirsch 1998 | Case report (n=1) |
| Hirsch 1999 | Case report (n=1) |
| Hirsche 1998 | Case report (n=1) |
| Hirsche 1999 | Case report (n=1) |
| Hixon & Oldfather 1958 | No incisor extraction |
| Hixon et al. 1969 | No incisor extraction |
| Hixon et al. 1971 | No incisor extraction |
| Hjermstad 1998 | Case report (n=1) |
| Hjermstad 1999 | Case report (n=1) |
| Ho 1998 | Case report (n=1) |
| Ho 1999 | Case report (n=1) |
| Hoang 1998 | Case report (n=1) |
| Hoang 1999 | Case report (n=1) |
| Hobson 1998 | Case report (n=1) |
| Hobson 1999 | Case report (n=1) |
| Hoch 1998 | Case report (n=1) |
| Hoch 1999 | Case report (n=1) |
| Hocking 1998 | Case report (n=1) |
| Hocking 1999 | Case report (n=1) |
| Hodge 1998 | Case report (n=1) |
| Hodge 1999 | Case report (n=1) |
| Hodgkins 1998 | Case report (n=1) |
| Hodgkins 1999 | Case report (n=1) |
| Hodgson 1998 | Case report (n=1) |
| Hodgson 1999 | Case report (n=1) |
| Hoediono 1998 | Case report (n=1) |
| Hoediono 1999 | Case report (n=1) |
| Hoefler 1998 | Case report (n=1) |
| Hoefler 1999 | Case report (n=1) |
| Hoekstra 1998 | Case report (n=1) |
| Hoekstra 1999 | Case report (n=1) |
| Hoelscher 1998 | Case report (n=1) |
| Hoelscher 1999 | Case report (n=1) |
| Hoerler 1998 | Case report (n=1) |
| Hoerler 1999 | Case report (n=1) |
| Hoerr 1998 | Case report (n=1) |
| Hoerr 1999 | Case report (n=1) |
| Hoerst 1998 | Case report (n=1) |
| Hoerst 1999 | Case report (n=1) |
| Hofer 1998 | Case report (n=1) |
| Hofer 1999 | Case report (n=1) |
| Hoff 1998 | Case report (n=1) |
| Hoff 1999 | Case report (n=1) |
| Hoffman 1998 | Case report (n=1) |
| Hoffman 1999 | Case report (n=1) |
| Hoffmann 1998 | Case report (n=1) |
| Hoffmann 1999 | Case report (n=1) |
| Hoffmeister 1998 | Case report (n=1) |
| Hoffmeister 1999 | Case report (n=1) |
| Hofmann 1998 | Case report (n=1) |
| Hofmann 1999 | Case report (n=1) |
| Hogan 1998 | Case report (n=1) |
| Hogan 1999 | Case report (n=1) |
| Hoggan 1998 | Case report (n=1) |
| Hoggan 1999 | Case report (n=1) |
| Hogue 1998 | Case report (n=1) |
| Hogue 1999 | Case report (n=1) |
| Hohenstein 1998 | Case report (n=1) |
| Hohenstein 1999 | Case report (n=1) |
| Hohlt 1998 | Case report (n=1) |
| Hohlt 1999 | Case report (n=1) |
| Hoj 1998 | Case report (n=1) |
| Hoj 1999 | Case report (n=1) |
| Holberg et al. 2005 | Finite element study |
| Holberg et al. 2007 | Finite element study |
| Holberg et al. 2012 | Finite element study |
| Holberg et al. 2013 | Finite element study |
| Holberg et al. 2014 | Finite element study |
| Holden 1998 | Case report (n=1) |
| Holden 1999 | Case report (n=1) |
| Holder 1998 | Case report (n=1) |
| Holder 1999 | Case report (n=1) |
| Holderman 1998 | Case report (n=1) |
| Holderman 1999 | Case report (n=1) |
| Holdsworth 1998 | Case report (n=1) |
| Holdsworth 1999 | Case report (n=1) |
| Hole 1998 | Case report (n=1) |
| Hole 1999 | Case report (n=1) |
| Holguin 1998 | Case report (n=1) |
| Holguin 1999 | Case report (n=1) |
| Holifield 1998 | Case report (n=1) |
| Holifield 1999 | Case report (n=1) |
| Holland 1998 | Case report (n=1) |
| Holland 1999 | Case report (n=1) |
| Holleman 1998 | Case report (n=1) |
| Holleman 1999 | Case report (n=1) |
| Hollender 1998 | Case report (n=1) |
| Hollender 1999 | Case report (n=1) |
| Holliday 1998 | Case report (n=1) |
| Holliday 1999 | Case report (n=1) |
| Hollingsworth 1998 | Case report (n=1) |
| Hollingsworth 1999 | Case report (n=1) |
| Hollis 1998 | Case report (n=1) |
| Hollis 1999 | Case report (n=1) |
| Hollister 1998 | Case report (n=1) |
| Hollister 1999 | Case report (n=1) |
| Holloway 1998 | Case report (n=1) |
| Holloway 1999 | Case report (n=1) |
| Holly 1998 | Case report (n=1) |
| Holly 1999 | Case report (n=1) |
| Holm 1998 | Case report (n=1) |
| Holm 1999 | Case report (n=1) |
| Holman 1998 | Case report (n=1) |
| Holman 1999 | Case report (n=1) |
| Holmes 1998 | Case report (n=1) |
| Holmes 1999 | Case report (n=1) |
| Holmgren 1998 | Case report (n=1) |
| Holmgren 1999 | Case report (n=1) |
| Holmquist 1998 | Case report (n=1) |
| Holmquist 1999 | Case report (n=1) |
| Holroyd 1998 | Case report (n=1) |
| Holroyd 1999 | Case report (n=1) |
| Holst 1998 | Case report (n=1) |
| Holst 1999 | Case report (n=1) |
| Holt 1998 | Case report (n=1) |
| Holt 1999 | Case report (n=1) |
| Holte 1998 | Case report (n=1) |
| Holte 1999 | Case report (n=1) |
| Holton 1998 | Case report (n=1) |
| Holton 1999 | Case report (n=1) |
| Holtz 1998 | Case report (n=1) |
| Holtz 1999 | Case report (n=1) |
| Holub 1998 | Case report (n=1) |
| Holub 1999 | Case report (n=1) |
| Holz 1998 | Case report (n=1) |
| Holz 1999 | Case report (n=1) |
| Holzapfel 1998 | Case report (n=1) |
| Holzapfel 1999 | Case report (n=1) |
| Homayoun 1998 | Case report (n=1) |
| Homayoun 1999 | Case report (n=1) |
| Hombach 1998 | Case report (n=1) |
| Hombach 1999 | Case report (n=1) |
| Homer 1998 | Case report (n=1) |
| Homer 1999 | Case report (n=1) |
| Hompesch 1998 | Case report (n=1) |
| Hompesch 1999 | Case report (n=1) |
| Honan 1998 | Case report (n=1) |
| Honan 1999 | Case report (n=1) |
| Honey 1998 | Case report (n=1) |
| Honey 1999 | Case report (n=1) |
| Honeyman 1998 | Case report (n=1) |
| Honeyman 1999 | Case report (n=1) |
| Hong 1998 | Case report (n=1) |
| Hong 1999 | Case report (n=1) |
| Hong et al. 2011 | Finite element study |
| Hong et al. 2015 | Animal study |
| Honigman 1998 | Case report (n=1) |
| Honigman 1999 | Case report (n=1) |
| Hönn & Göz 2007 | Review article |
| Hönn et al. 2009 | Finite element study |
| Hood 1998 | Case report (n=1) |
| Hood 1999 | Case report (n=1) |
| Hoogeveen 1998 | Case report (n=1) |
| Hoogeveen 1999 | Case report (n=1) |
| Hook 1998 | Case report (n=1) |
| Hook 1999 | Case report (n=1) |
| Hooper 1998 | Case report (n=1) |
| Hooper 1999 | Case report (n=1) |
| Hooten 1998 | Case report (n=1) |
| Hooten 1999 | Case report (n=1) |
| Hoover 1998 | Case report (n=1) |
| Hoover 1999 | Case report (n=1) |
| Hope 1998 | Case report (n=1) |
| Hope 1999 | Case report (n=1) |
| Hopkin 1998 | Case report (n=1) |
| Hopkin 1999 | Case report (n=1) |
| Hopkins 1998 | Case report (n=1) |
| Hopkins 1999 | Case report (n=1) |
| Hoppe 1998 | Case report (n=1) |
| Hoppe 1999 | Case report (n=1) |
| Hoppenreijs et al. 1998 | No incisor extraction |
| Hoppenreijs et al. 1999 | No incisor extraction |
| Hopps 1998 | Case report (n=1) |
| Hopps 1999 | Case report (n=1) |
| Horgan 1998 | Case report (n=1) |
| Horgan 1999 | Case report (n=1) |
| Hori 1998 | Case report (n=1) |
| Hori 1999 | Case report (n=1) |
| Horiuchi et al. 1998 | No incisor extraction |
| Horiuchi et al. 1999 | No incisor extraction |
| Horman 1998 | Case report (n=1) |
| Horman 1999 | Case report (n=1) |
| Horn 1998 | Case report (n=1) |
| Horn 1999 | Case report (n=1) |
| Horn et al. 2014 | Finite element study |
| Hornbuckle 1998 | Case report (n=1) |
| Hornbuckle 1999 | Case report (n=1) |
| Horne 1998 | Case report (n=1) |
| Horne 1999 | Case report (n=1) |
| Horner 1998 | Case report (n=1) |
| Horner 1999 | Case report (n=1) |
| Hornsby 1998 | Case report (n=1) |
| Hornsby 1999 | Case report (n=1) |
| Horowitz 1998 | Case report (n=1) |
| Horowitz 1999 | Case report (n=1) |
| Horrocks 1998 | Case report (n=1) |
| Horrocks 1999 | Case report (n=1) |
| Horsley 1998 | Case report (n=1) |
| Horsley 1999 | Case report (n=1) |
| Horst 1998 | Case report (n=1) |
| Horst 1999 | Case report (n=1) |
| Horton 1998 | Case report (n=1) |
| Horton 1999 | Case report (n=1) |
| Horvath 1998 | Case report (n=1) |
| Horvath 1999 | Case report (n=1) |
| Horwitz 1998 | Case report (n=1) |
| Horwitz 1999 | Case report (n=1) |
| Hosfield 1998 | Case report (n=1) |
| Hosfield 1999 | Case report (n=1) |
| Hosgood 1998 | Case report (n=1) |
| Hosgood 1999 | Case report (n=1) |
| Hoskens 1998 | Case report (n=1) |
| Hoskens 1999 | Case report (n=1) |
| Hosking 1998 | Case report (n=1) |
| Hosking 1999 | Case report (n=1) |
| Hoskinson 1998 | Case report (n=1) |
| Hoskinson 1999 | Case report (n=1) |
| Hosmer 1998 | Case report (n=1) |
| Hosmer 1999 | Case report (n=1) |
| Hoss 1998 | Case report (n=1) |
| Hoss 1999 | Case report (n=1) |
| Hosseini 1998 | Case report (n=1) |
| Hosseini 1999 | Case report (n=1) |
| Host 1998 | Case report (n=1) |
| Host 1999 | Case report (n=1) |
| Hotchkiss 1998 | Case report (n=1) |
| Hotchkiss 1999 | Case report (n=1) |
| Hoth 1998 | Case report (n=1) |
| Hoth 1999 | Case report (n=1) |
| Hottel 1998 | Case report (n=1) |
| Hottel 1999 | Case report (n=1) |
| Hotz 1998 | Case report (n=1) |
| Hotz 1999 | Case report (n=1) |
| Hou 1998 | Case report (n=1) |
| Hou 1999 | Case report (n=1) |
| Houghton 1998 | Case report (n=1) |
| Houghton 1999 | Case report (n=1) |
| Houlden 1998 | Case report (n=1) |
| Houlden 1999 | Case report (n=1) |
| House 1998 | Case report (n=1) |
| House 1999 | Case report (n=1) |
| Houser 1998 | Case report (n=1) |
| Houser 1999 | Case report (n=1) |
| Houston 1998 | Case report (n=1) |
| Houston 1999 | Case report (n=1) |
| Hovell 1998 | Case report (n=1) |
| Hovell 1999 | Case report (n=1) |
| Hoven 1998 | Case report (n=1) |
| Hoven 1999 | Case report (n=1) |
| Hovis 1998 | Case report (n=1) |
| Hovis 1999 | Case report (n=1) |
| Howard 1998 | Case report (n=1) |
| Howard 1999 | Case report (n=1) |
| Howat 1998 | Case report (n=1) |
| Howat 1999 | Case report (n=1) |
| Howe 1998 | Case report (n=1) |
| Howe 1999 | Case report (n=1) |
| Howell 1998 | Case report (n=1) |
| Howell 1999 | Case report (n=1) |
| Howells 1998 | Case report (n=1) |
| Howells 1999 | Case report (n=1) |
| Howes 1998 | Case report (n=1) |
| Howes 1999 | Case report (n=1) |
| Howett 1998 | Case report (n=1) |
| Howett 1999 | Case report (n=1) |
| Howie 1998 | Case report (n=1) |
| Howie 1999 | Case report (n=1) |
| Howitt 1998 | Case report (n=1) |
| Howitt 1999 | Case report (n=1) |
| Howker 1998 | Case report (n=1) |
| Howker 1999 | Case report (n=1) |
| Howland 1998 | Case report (n=1) |
| Howland 1999 | Case report (n=1) |
| Howlett 1998 | Case report (n=1) |
| Howlett 1999 | Case report (n=1) |
| Howley 1998 | Case report (n=1) |
| Howley 1999 | Case report (n=1) |
| Hoxie 1998 | Case report (n=1) |
| Hoxie 1999 | Case report (n=1) |
| Hoy 1998 | Case report (n=1) |
| Hoy 1999 | Case report (n=1) |
| Hoybjerg 1998 | Case report (n=1) |
| Hoybjerg 1999 | Case report (n=1) |
| Hoyer 1998 | Case report (n=1) |
| Hoyer 1999 | Case report (n=1) |
| Hoyle 1998 | Case report (n=1) |
| Hoyle 1999 | Case report (n=1) |
| Hoyt 1998 | Case report (n=1) |
| Hoyt 1999 | Case report (n=1) |
| Hrivnak 1998 | Case report (n=1) |
| Hrivnak 1999 | Case report (n=1) |
| Hronek 1998 | Case report (n=1) |
| Hronek 1999 | Case report (n=1) |
| Hruby 1998 | Case report (n=1) |
| Hruby 1999 | Case report (n=1) |
| Hruska 1998 | Case report (n=1) |
| Hruska 1999 | Case report (n=1) |
| Hrycyshyn 1998 | Case report (n=1) |
| Hrycyshyn 1999 | Case report (n=1) |
| Hsu 1998 | Case report (n=1) |
| Hsu 1999 | Case report (n=1) |
| Hsueh 1998 | Case report (n=1) |
| Hsueh 1999 | Case report (n=1) |
| Hu 1998 | Case report (n=1) |
| Hu 1999 | Case report (n=1) |
| Huang 1998 | Case report (n=1) |
| Huang 1999 | Case report (n=1) |
| Huang et al. 2005 | Finite element study |
| Huang et al. 2012 | Finite element study |
| Huang et al. 2014 | Animal study |
| Huang et al. 2018 | No quantitative data |
| Hubbard 1998 | Case report (n=1) |
| Hubbard 1999 | Case report (n=1) |
| Huber 1998 | Case report (n=1) |
| Huber 1999 | Case report (n=1) |
| Hubert 1998 | Case report (n=1) |
| Hubert 1999 | Case report (n=1) |
| Hubler 1998 | Case report (n=1) |
| Hubler 1999 | Case report (n=1) |
| Huddle 1998 | Case report (n=1) |
| Huddle 1999 | Case report (n=1) |
| Huddleston 1998 | Case report (n=1) |
| Huddleston 1999 | Case report (n=1) |
| Hudec 1998 | Case report (n=1) |
| Hudec 1999 | Case report (n=1) |
| Hudgins 1998 | Case report (n=1) |
| Hudgins 1999 | Case report (n=1) |
| Hudson 1998 | Case report (n=1) |
| Hudson 1999 | Case report (n=1) |
| Huerta 1998 | Case report (n=1) |
| Huerta 1999 | Case report (n=1) |
| Huff 1998 | Case report (n=1) |
| Huff 1999 | Case report (n=1) |
| Huffman 1998 | Case report (n=1) |
| Huffman 1999 | Case report (n=1) |
| Hufnagel 1998 | Case report (n=1) |
| Hufnagel 1999 | Case report (n=1) |
| Hug 1998 | Case report (n=1) |
| Hug 1999 | Case report (n=1) |
| Hugg 1998 | Case report (n=1) |
| Hugg 1999 | Case report (n=1) |
| Huggins 1998 | Case report (n=1) |
| Huggins 1999 | Case report (n=1) |
| Hughes 1998 | Case report (n=1) |
| Hughes 1999 | Case report (n=1) |
| Hughes et al. 2014 | No incisor extraction |
| Hughey 1998 | Case report (n=1) |
| Hughey 1999 | Case report (n=1) |
| Hugly 1998 | Case report (n=1) |
| Hugly 1999 | Case report (n=1) |
| Hugo 1998 | Case report (n=1) |
| Hugo 1999 | Case report (n=1) |
| Huguet 1998 | Case report (n=1) |
| Huguet 1999 | Case report (n=1) |
| Huh 1998 | Case report (n=1) |
| Huh 1999 | Case report (n=1) |
| Hujer 1998 | Case report (n=1) |
| Hujer 1999 | Case report (n=1) |
| Hulen 1998 | Case report (n=1) |
| Hulen 1999 | Case report (n=1) |
| Huling 1998 | Case report (n=1) |
| Huling 1999 | Case report (n=1) |
| Hulk 1998 | Case report (n=1) |
| Hulk 1999 | Case report (n=1) |
| Hull 1998 | Case report (n=1) |
| Hull 1999 | Case report (n=1) |
| Huls 1998 | Case report (n=1) |
| Huls 1999 | Case report (n=1) |
| Hulsey 1998 | Case report (n=1) |
| Hulsey 1999 | Case report (n=1) |
| Hult 1998 | Case report (n=1) |
| Hult 1999 | Case report (n=1) |
| Hultberg 1998 | Case report (n=1) |
| Hultberg 1999 | Case report (n=1) |
| Hultgren 1998 | Case report (n=1) |
| Hultgren 1999 | Case report (n=1) |
| Hultman 1998 | Case report (n=1) |
| Hultman 1999 | Case report (n=1) |
| Hultz 1998 | Case report (n=1) |
| Hultz 1999 | Case report (n=1) |
| Humber 1998 | Case report (n=1) |
| Humber 1999 | Case report (n=1) |
| Humbert 1998 | Case report (n=1) |
| Humbert 1999 | Case report (n=1) |
| Humbles 1998 | Case report (n=1) |
| Humbles 1999 | Case report (n=1) |
| Humes 1998 | Case report (n=1) |
| Humes 1999 | Case report (n=1) |
| Humm 1998 | Case report (n=1) |
| Humm 1999 | Case report (n=1) |
| Hummel 1998 | Case report (n=1) |
| Hummel 1999 | Case report (n=1) |
| Humphrey 1998 | Case report (n=1) |
| Humphrey 1999 | Case report (n=1) |
| Humphreys 1998 | Case report (n=1) |
| Humphreys 1999 | Case report (n=1) |
| Humphries 1998 | Case report (n=1) |
| Humphries 1999 | Case report (n=1) |
| Hung 1998 | Case report (n=1) |
| Hung 1999 | Case report (n=1) |
| Hungerford 1998 | Case report (n=1) |
| Hungerford 1999 | Case report (n=1) |
| Hunkins 1998 | Case report (n=1) |
| Hunkins 1999 | Case report (n=1) |
| Hunsaker 1998 | Case report (n=1) |
| Hunsaker 1999 | Case report (n=1) |
| Hunt 1998 | Case report (n=1) |
| Hunt 1999 | Case report (n=1) |
| Hunter 1998 | Case report (n=1) |
| Hunter 1999 | Case report (n=1) |
| Hunter et al. 2007 | No incisor extraction |
| Hunting 1998 | Case report (n=1) |
| Hunting 1999 | Case report (n=1) |
| Huntington 1998 | Case report (n=1) |
| Huntington 1999 | Case report (n=1) |
| Huntley 1998 | Case report (n=1) |
| Huntley 1999 | Case report (n=1) |
| Hunziker 1998 | Case report (n=1) |
| Hunziker 1999 | Case report (n=1) |
| Hupf 1998 | Case report (n=1) |
| Hupf 1999 | Case report (n=1) |
| Hupp 1998 | Case report (n=1) |
| Hupp 1999 | Case report (n=1) |
| Hurd 1998 | Case report (n=1) |
| Hurd 1999 | Case report (n=1) |
| Hurley 1998 | Case report (n=1) |
| Hurley 1999 | Case report (n=1) |
| Hurmerinta 1998 | Case report (n=1) |
| Hurmerinta 1999 | Case report (n=1) |
| Hurmuz 1998 | Case report (n=1) |
| Hurmuz 1999 | Case report (n=1) |
| Hurn 1998 | Case report (n=1) |
| Hurn 1999 | Case report (n=1) |
| Hurst 1998 | Case report (n=1) |
| Hurst 1999 | Case report (n=1) |
| Hurt 1998 | Case report (n=1) |
| Hurt 1999 | Case report (n=1) |
| Hurwitz 1998 | Case report (n=1) |
| Hurwitz 1999 | Case report (n=1) |
| Husain 1998 | Case report (n=1) |
| Husain 1999 | Case report (n=1) |
| Husak 1998 | Case report (n=1) |
| Husak 1999 | Case report (n=1) |
| Husbands 1998 | Case report (n=1) |
| Husbands 1999 | Case report (n=1) |
| Huse 1998 | Case report (n=1) |
| Huse 1999 | Case report (n=1) |
| Huseman 1998 | Case report (n=1) |
| Huseman 1999 | Case report (n=1) |
| Huser 1998 | Case report (n=1) |
| Huser 1999 | Case report (n=1) |
| Huston 1998 | Case report (n=1) |
| Huston 1999 | Case report (n=1) |
| Hutchens 1998 | Case report (n=1) |
| Hutchens 1999 | Case report (n=1) |
| Hutchings 1998 | Case report (n=1) |
| Hutchings 1999 | Case report (n=1) |
| Hutchins 1998 | Case report (n=1) |
| Hutchins 1999 | Case report (n=1) |
| Hutchinson 1998 | Case report (n=1) |
| Hutchinson 1999 | Case report (n=1) |
| Hutchison 1998 | Case report (n=1) |
| Hutchison 1999 | Case report (n=1) |
| Huth 1998 | Case report (n=1) |
| Huth 1999 | Case report (n=1) |
| Hutson 1998 | Case report (n=1) |
| Hutson 1999 | Case report (n=1) |
| Hutt 1998 | Case report (n=1) |
| Hutt 1999 | Case report (n=1) |
| Hutten 1998 | Case report (n=1) |
| Hutten 1999 | Case report (n=1) |
| Hutter 1998 | Case report (n=1) |
| Hutter 1999 | Case report (n=1) |
| Hutton 1998 | Case report (n=1) |
| Hutton 1999 | Case report (n=1) |
| Hutz 1998 | Case report (n=1) |
| Hutz 1999 | Case report (n=1) |
| Hux 1998 | Case report (n=1) |
| Hux 1999 | Case report (n=1) |
| Huxley 1998 | Case report (n=1) |
| Huxley 1999 | Case report (n=1) |
| Huxtable 1998 | Case report (n=1) |
| Huxtable 1999 | Case report (n=1) |
| Huy 1998 | Case report (n=1) |
| Huy 1999 | Case report (n=1) |
| Huynh 1998 | Case report (n=1) |
| Huynh 1999 | Case report (n=1) |
| Hwang 1998 | Case report (n=1) |
| Hwang 1999 | Case report (n=1) |
| Hwang et al. 2014 | Finite element study |
| Hwang et al. 2017 | Animal study |
| Hwu 1998 | Case report (n=1) |
| Hwu 1999 | Case report (n=1) |
| Hyatt 1998 | Case report (n=1) |
| Hyatt 1999 | Case report (n=1) |
| Hyde 1998 | Case report (n=1) |
| Hyde 1999 | Case report (n=1) |
| Hyder 1998 | Case report (n=1) |
| Hyder 1999 | Case report (n=1) |
| Hyer 1998 | Case report (n=1) |
| Hyer 1999 | Case report (n=1) |
| Hyland 1998 | Case report (n=1) |
| Hyland 1999 | Case report (n=1) |
| Hylton 1998 | Case report (n=1) |
| Hylton 1999 | Case report (n=1) |
| Hyman 1998 | Case report (n=1) |
| Hyman 1999 | Case report (n=1) |
| Hymes 1998 | Case report (n=1) |
| Hymes 1999 | Case report (n=1) |
| Hymowitz 1998 | Case report (n=1) |
| Hymowitz 1999 | Case report (n=1) |
| Hynd 1998 | Case report (n=1) |
| Hynd 1999 | Case report (n=1) |
| Hynek 1998 | Case report (n=1) |
| Hynek 1999 | Case report (n=1) |
| Hynes 1998 | Case report (n=1) |
| Hynes 1999 | Case report (n=1) |
| Hysell 1998 | Case report (n=1) |
| Hysell 1999 | Case report (n=1) |
| Hyslop 1998 | Case report (n=1) |
| Hyslop 1999 | Case report (n=1) |
| Hytönen 1998 | Case report (n=1) |
| Hytönen 1999 | Case report (n=1) |
| Hyvärinen 1998 | Case report (n=1) |
| Hyvärinen 1999 | Case report (n=1) |
| Hyvönen 1998 | Case report (n=1) |
| Hyvönen 1999 | Case report (n=1) |
| Iacob 1998 | Case report (n=1) |
| Iacob 1999 | Case report (n=1) |
| Iacono 1998 | Case report (n=1) |
| Iacono 1999 | Case report (n=1) |
| Iacopino 1998 | Case report (n=1) |
| Iacopino 1999 | Case report (n=1) |
| Iacovelli 1998 | Case report (n=1) |
| Iacovelli 1999 | Case report (n=1) |
| Iacoviello 1998 | Case report (n=1) |
| Iacoviello 1999 | Case report (n=1) |
| Iacovino 1998 | Case report (n=1) |
| Iacovino 1999 | Case report (n=1) |
| Iacovitti 1998 | Case report (n=1) |
| Iacovitti 1999 | Case report (n=1) |
| Iacullo 1998 | Case report (n=1) |
| Iacullo 1999 | Case report (n=1) |
| Iadipaolo 1998 | Case report (n=1) |
| Iadipaolo 1999 | Case report (n=1) |
| Iafrate 1998 | Case report (n=1) |
| Iafrate 1999 | Case report (n=1) |
| Iafrati 1998 | Case report (n=1) |
| Iafrati 1999 | Case report (n=1) |
| Iafusco 1998 | Case report (n=1) |
| Iafusco 1999 | Case report (n=1) |
| Iagnocco 1998 | Case report (n=1) |
| Iagnocco 1999 | Case report (n=1) |
| Iagrossi 1998 | Case report (n=1) |
| Iagrossi 1999 | Case report (n=1) |
| Iagulli 1998 | Case report (n=1) |
| Iagulli 1999 | Case report (n=1) |
| Ialeggio 1998 | Case report (n=1) |
| Ialeggio 1999 | Case report (n=1) |
| Ialenti 1998 | Case report (n=1) |
| Ialenti 1999 | Case report (n=1) |
| Ialongo 1998 | Case report (n=1) |
| Ialongo 1999 | Case report (n=1) |
| Iamacone 1998 | Case report (n=1) |
| Iamacone 1999 | Case report (n=1) |
| Iamadrid 1998 | Case report (n=1) |
| Iamadrid 1999 | Case report (n=1) |
| Iamaf 1998 | Case report (n=1) |
| Iamaf 1999 | Case report (n=1) |
| Iamai 1998 | Case report (n=1) |
| Iamai 1999 | Case report (n=1) |
| Iamale 1998 | Case report (n=1) |
| Iamale 1999 | Case report (n=1) |
| Iaman 1998 | Case report (n=1) |
| Iaman 1999 | Case report (n=1) |
| Iamandi 1998 | Case report (n=1) |
| Iamandi 1999 | Case report (n=1) |
| Iandoli 1998 | Case report (n=1) |
| Iandoli 1999 | Case report (n=1) |
| Iandolo 1998 | Case report (n=1) |
| Iandolo 1999 | Case report (n=1) |
| Iandiorio 1998 | Case report (n=1) |
| Iandiorio 1999 | Case report (n=1) |
| Iandoli 1998 | Case report (n=1) |
| Iandoli 1999 | Case report (n=1) |
| Iandolo 1998 | Case report (n=1) |
| Iandolo 1999 | Case report (n=1) |
| Iandria 1998 | Case report (n=1) |
| Iandria 1999 | Case report (n=1) |
| Iandriello 1998 | Case report (n=1) |
| Iandriello 1999 | Case report (n=1) |
| Iandrio 1998 | Case report (n=1) |
| Iandrio 1999 | Case report (n=1) |
| Iandrs 1998 | Case report (n=1) |
| Iandrs 1999 | Case report (n=1) |
| Iandu 1998 | Case report (n=1) |
| Iandu 1999 | Case report (n=1) |
| Ianduca 1998 | Case report (n=1) |
| Ianduca 1999 | Case report (n=1) |
| Iandur 1998 | Case report (n=1) |
| Iandur 1999 | Case report (n=1) |
| Iandus 1998 | Case report (n=1) |
| Iandus 1999 | Case report (n=1) |
| Iandv 1998 | Case report (n=1) |
| Iandv 1999 | Case report (n=1) |
| Iandw 1998 | Case report (n=1) |
| Iandw 1999 | Case report (n=1) |
| Iandx 1998 | Case report (n=1) |
| Iandx 1999 | Case report (n=1) |
| Iandy 1998 | Case report (n=1) |
| Iandy 1999 | Case report (n=1) |
| Iandz 1998 | Case report (n=1) |
| Iandz 1999 | Case report (n=1) |
| Iane 1998 | Case report (n=1) |
| Iane 1999 | Case report (n=1) |
| Ianecko 1998 | Case report (n=1) |
| Ianecko 1999 | Case report (n=1) |
| Ianeira 1998 | Case report (n=1) |
| Ianeira 1999 | Case report (n=1) |
| Ianeiro 1998 | Case report (n=1) |
| Ianeiro 1999 | Case report (n=1) |
| Ianeirot 1998 | Case report (n=1) |
| Ianeirot 1999 | Case report (n=1) |
| Ianeirov 1998 | Case report (n=1) |
| Ianeirov 1999 | Case report (n=1) |
| Ianeirow 1998 | Case report (n=1) |
| Ianeirow 1999 | Case report (n=1) |
| Ianeirp 1998 | Case report (n=1) |
| Ianeirp 1999 | Case report (n=1) |
| Ianeirs 1998 | Case report (n=1) |
| Ianeirs 1999 | Case report (n=1) |
| Ianeirt 1998 | Case report (n=1) |
| Ianeirt 1999 | Case report (n=1) |
| Ianeiru 1998 | Case report (n=1) |
| Ianeiru 1999 | Case report (n=1) |
| Ianeirv 1998 | Case report (n=1) |
| Ianeirv 1999 | Case report (n=1) |
| Ianeirw 1998 | Case report (n=1) |
| Ianeirw 1999 | Case report (n=1) |
| Ianeirx 1998 | Case report (n=1) |
| Ianeirx 1999 | Case report (n=1) |
| Ianeiry 1998 | Case report (n=1) |
| Ianeiry 1999 | Case report (n=1) |
| Ianeirz 1998 | Case report (n=1) |
| Ianeirz 1999 | Case report (n=1) |
| Ianeis 1998 | Case report (n=1) |
| Ianeis 1999 | Case report (n=1) |
| Ianeit 1998 | Case report (n=1) |
| Ianeit 1999 | Case report (n=1) |
| Ianeiu 1998 | Case report (n=1) |
| Ianeiu 1999 | Case report (n=1) |
| Ianeiv 1998 | Case report (n=1) |
| Ianeiv 1999 | Case report (n=1) |
| Ianeiw 1998 | Case report (n=1) |
| Ianeiw 1999 | Case report (n=1) |
| Ianeix 1998 | Case report (n=1) |
| Ianeix 1999 | Case report (n=1) |
| Ianeiy 1998 | Case report (n=1) |
| Ianeiy 1999 | Case report (n=1) |
| Ianeiz 1998 | Case report (n=1) |
| Ianeiz 1999 | Case report (n=1) |
| Ianeja 1998 | Case report (n=1) |
| Ianeja 1999 | Case report (n=1) |
| Ianejac 1998 | Case report (n=1) |
| Ianejac 1999 | Case report (n=1) |
| Ianejag 1998 | Case report (n=1) |
| Ianejag 1999 | Case report (n=1) |
| Ianejak 1998 | Case report (n=1) |
| Ianejak 1999 | Case report (n=1) |
| Ianejal 1998 | Case report (n=1) |
| Ianejal 1999 | Case report (n=1) |
| Ianejam 1998 | Case report (n=1) |
| Ianejam 1999 | Case report (n=1) |
| Ianejan 1998 | Case report (n=1) |
| Ianejan 1999 | Case report (n=1) |
| Ianejao 1998 | Case report (n=1) |
| Ianejao 1999 | Case report (n=1) |
| Ianejap 1998 | Case report (n=1) |
| Ianejap 1999 | Case report (n=1) |
| Ianejaq 1998 | Case report (n=1) |
| Ianejaq 1999 | Case report (n=1) |
| Ianejar 1998 | Case report (n=1) |
| Ianejar 1999 | Case report (n=1) |
| Ianejas 1998 | Case report (n=1) |
| Ianejas 1999 | Case report (n=1) |
| Ianejat 1998 | Case report (n=1) |
| Ianejat 1999 | Case report (n=1) |
| Ianejau 1998 | Case report (n=1) |
| Ianejau 1999 | Case report (n=1) |
| Ianejav 1998 | Case report (n=1) |
| Ianejav 1999 | Case report (n=1) |
| Ianejaw 1998 | Case report (n=1) |
| Ianejaw 1999 | Case report (n=1) |
| Ianejax 1998 | Case report (n=1) |
| Ianejax 1999 | Case report (n=1) |
| Ianejay 1998 | Case report (n=1) |
| Ianejay 1999 | Case report (n=1) |
| Ianejaz 1998 | Case report (n=1) |
| Ianejaz 1999 | Case report (n=1) |
| Ianejba 1998 | Case report (n=1) |
| Ianejba 1999 | Case report (n=1) |
| Ianejbc 1998 | Case report (n=1) |
| Ianejbc 1999 | Case report (n=1) |
| Ianejbd 1998 | Case report (n=1) |
| Ianejbd 1999 | Case report (n=1) |
| Ianejbe 1998 | Case report (n=1) |
| Ianejbe 1999 | Case report (n=1) |
| Ianejbf 1998 | Case report (n=1) |
| Ianejbf 1999 | Case report (n=1) |
| Ianejbg 1998 | Case report (n=1) |
| Ianejbg 1999 | Case report (n=1) |
| Ianejbh 1998 | Case report (n=1) |
| Ianejbh 1999 | Case report (n=1) |
| Ianejbi 1998 | Case report (n=1) |
| Ianejbi 1999 | Case report (n=1) |
| Ianejbj 1998 | Case report (n=1) |
| Ianejbj 1999 | Case report (n=1) |
| Iejbk 1998 | Case report (n=1) |
| Iejbk 1999 | Case report (n=1) |
| Iejbl 1998 | Case report (n=1) |
| Iejbl 1999 | Case report (n=1) |
| Iejbm 1998 | Case report (n=1) |
| Iejbm 1999 | Case report (n=1) |
| Iejbn 1998 | Case report (n=1) |
| Iejbn 1999 | Case report (n=1) |
| Iejbo 1998 | Case report (n=1) |
| Iejbo 1999 | Case report (n=1) |
| Iejbp 1998 | Case report (n=1) |
| Iejbp 1999 | Case report (n=1) |
| Iejbq 1998 | Case report (n=1) |
| Iejbq 1999 | Case report (n=1) |
| Iejbr 1998 | Case report (n=1) |
| Iejbr 1999 | Case report (n=1) |
| Iejbs 1998 | Case report (n=1) |
| Iejbs 1999 | Case report (n=1) |
| Iejbt 1998 | Case report (n=1) |
| Iejbt 1999 | Case report (n=1) |
| Iejbu 1998 | Case report (n=1) |
| Iejbu 1999 | Case report (n=1) |
| Iejbv 1998 | Case report (n=1) |
| Iejbv 1999 | Case report (n=1) |
| Iejbw 1998 | Case report (n=1) |
| Iejbw 1999 | Case report (n=1) |
| Iejbx 1998 | Case report (n=1) |
| Iejbx 1999 | Case report (n=1) |
| Iejby 1998 | Case report (n=1) |
| Iejby 1999 | Case report (n=1) |
| Iejbz 1998 | Case report (n=1) |
| Iejbz 1999 | Case report (n=1) |
